# Polygenic risk-by-socio-economic status interaction effects on specific and aggregated outcomes of depression, anxiety, body mass index, waist-hip-ratio, smoking and alcohol use

**DOI:** 10.1101/2025.09.22.25336352

**Authors:** Rujia Wang, Lifelines Cohort Study, Harold Snieder, Catharina A. Hartman

## Abstract

**Background:** Psychiatric and somatic problems are highly prevalent, often co-occur and share part of their genetic background. Low socio-economic status (SES) is a risk factor for both. We examined whether low SES amplifies the effects of genetic susceptibility on depression, anxiety, body mass index (BMI), waist-hip-ratio (WHR), smoking and alcohol use, investigating and aggregated outcomes.

**Methods:** Data came from the population-based Lifelines Cohort Study (n=50,761). Anxiety, depression, body mass index, waist-hip-ratio, smoking, and alcohol use were analyzed individually and collectively using confirmatory factor analysis to model shared variance among outcomes. Polygenic risk scores (PRSs) were calculated based on recent genome-wide association studies (GWASs) of specific conditions. We performed GWASs and derived PRS of aggregated outcomes using genomic structural equation modelling (SEM). A latent SES factor was generated by educational attainment, occupational status, and disposable household income. Interaction effects of SES indices with PRSs were estimated using linear mixed regression.

**Results:** Eight of ten PRSs-by-SES interactions were significant (*ps*<0.05) for depression, anxiety, BMI, smoking, and aggregated outcome level, but not for WHR (*p*=0.07) and alcohol use (*p*=0.67). Lower SES amplified the effects of PRSs on depression, anxiety, BMI, and smoking. At the aggregated outcome level, interaction effects were mostly smaller than effects for individual outcomes.

**Conclusions:** Our study demonstrates polygenic risk-by-SES interaction effects on depression, anxiety, BMI, and smoking. However, the effects were attenuated for PRSs derived from genomic SEM and aggregated outcomes, indicating limited additional etiological insights from modelling genetic and phenotypic overlap.

## Introduction

Depression, anxiety, obesity and substance use disorders are common diseases, and a major cause of long-term disability and mortality worldwide (Nigatu et al., 2016, Vos et al., 2016). These diseases are heritable, with estimated heritabilities in family and twin studies of 0.37 for depression (Sullivan et al., 2000), 0.32 for generalized anxiety disorder (Hettema et al., 2001), 0.46 for body mass index (BMI) (Elks et al., 2012), 0.30 for waist-hip-ratio (WHR) (Wang et al., 2022), and 0.57 and 0.67 for alcohol dependence and nicotine dependence (Sullivan et al., 2012), respectively. As depression, anxiety, obesity and substance use are polygenic diseases, polygenic risk scores (PRS) can be used to capture genetic susceptibility based on common genetic variants identified in genome-wide association studies (GWASs).

In addition to genetic effects, gradients in socio-economic status (SES) show strong associations with depression, anxiety, obesity and substance use, with lower SES groups experiencing poorer mental and physical health (Cohen et al., 2020, Gu & Ming, 2020, Mohammed et al., 2019, Park et al., 2020, Pennanen et al., 2014, Reid et al., 2010, Richardson et al., 2015, Valencia et al., 2019, Yoo et al., 2016). SES is a complex and multifactorial construct, encompassing individual, household, and neighborhood levels (Braveman et al., 2005). Individuals with lower educational attainment (EA) levels were more likely to experience depression (Cohen et al., 2020), obesity (Park et al., 2020), and were prone to alcoholism (Gu & Ming, 2020), and smoking (Reid et al., 2010) compared to those with higher educational attainment. Individuals with a lower occupational status were more likely to have depression (Yoo et al., 2016). Lower household income is associated with obesity (female) (Park et al., 2020), early initiation of smoking and heavy smoking (Pennanen et al., 2014, Valencia et al., 2019), while higher household income has been associated with alcoholism (Gu & Ming, 2020). Finally, lower neighborhood socio-economic status (NSES) has been associated with higher depression risk (Richardson et al., 2015), and obesity (Mohammed et al., 2019). In this study, we included educational attainment and occupational status for individual SES, disposable household income for household SES, and NSES.

Although both genetics and SES indices are associated with depression, anxiety, obesity and substance use, it is unknown to what extent low SES may moderate genetic effects on these outcomes. Few studies have explored the interplay between PRSs and SES. One study showed that lower parental SES amplified PRS effects on early-onset depression (Agerbo et al., 2021). For obesity, one found that SES during childhood moderated PRS effects on BMI, but only in one subgroup (white women) (Thompson et al., 2020), another that lower educational attainment amplified PRS of BMI effects on obesity, but not on BMI (Barcellos et al., 2018). Higher neighborhood income amplified a PRS effect for alcohol use but not smoking (Pasman et al., 2020). No previous studies have explored PRS-SES interaction for anxiety, likely due to previously underpowered GWAS.

Low SES is associated with mental and somatic conditions (Cohen et al., 2020, Gu & Ming, 2020, Mohammed et al., 2019, Park et al., 2020, Pennanen et al., 2014, Reid et al., 2010, Richardson et al., 2015, Valencia et al., 2019, Yoo et al., 2016), while depression and anxiety, obesity and substance use often co-occur (Nigatu et al., 2016). This raises the question if effects of SES are similar at this aggregated level, which can be modelled as the shared variance among these outcomes, by means of factor analysis. These conditions also co-aggregate within families and share genetic background (Anttila et al., 2018, Wang et al., 2022). For instance, genetic correlations (rG) are 0.94 between depression and anxiety, 0.54 between BMI and WHR, and 0.31 between number of cigarettes per day and daily alcohol intake (Wang et al., 2022), indicating shared genetic susceptibility may contribute to their phenotypic co-occurrence (Nigatu et al., 2016). Additionally, Milaneschi and colleagues found that genes near BMI-associated loci were highly expressed in specific brain regions that regulate both appetite and energy homeostasis and mood (Anttila et al., 2018). Furthermore, it may even be the case that shared genetic susceptibility may partly explain the shared variance across three domains of depression/anxiety, obesity, and substance use, with genetic correlations ranging from 0.16 to 0.39 (Anttila et al., 2018, Wang et al., 2022). Likewise, this raises the question of whether SES may interact with genetic susceptibility at this higher aggregated level. While genomic structural equation modelling (genomic SEM) has been used to explore shared etiology across psychiatric disorders in recent years (Grotzinger et al., 2022, Grotzinger et al., 2019, Lee et al., 2019, Levey et al., 2021), but it has not been applied to study the potential shared G×E mechanisms across conditions, which will be pursued here.

In Lifelines cohort study (N=50,761), we calculated PRSs for individual outcomes based on recent large GWASs (Howard et al., 2019, Liu et al., 2019, Pulit et al., 2019, Purves et al., 2020) and derived shared PRSs using genomic SEM (Grotzinger et al., 2019) to reflect shared genetic susceptibility between disease outcomes. Our aims were to investigate whether a latent SES factor as well as individual, household, or neighborhood SES, 1) moderated genetic susceptibility to individual outcomes representing depression and anxiety, BMI, WHR, smoking and alcohol use; 2) moderated shared genetic susceptibility to aggregated factors of depression and anxiety, BMI and WHR, and smoking and alcohol use.

## Methods

### Study sample and design

We used data from the ongoing Lifelines Cohort study, a multi-disciplinary prospective population-based study in the Netherlands, involving 167,729 participants across three generations. It employs a broad range of investigative procedures in assessing the biomedical, socio-demographic, behavioural, physical and psychological factors which contribute to the health and disease of the general population, with a special focus on multi-morbidity and complex genetics (Sijtsma et al., 2021). Baseline assessment took place between 2006 and 2013, with follow-up from 2014 to 2017. Genetic data is available for over 50,000 participants. The Lifelines Cohort study follows the principles of the Declaration of Helsinki and in accordance with the research code of University Medical Center Groningen, and the Lifelines protocol was approved by the UMCG Medical ethical committee (2007/152). All participants signed an informed consent form.

### Measurement Outcomes

Current depression and anxiety were measured using the MINI International Neuropsychiatric Interview (MINI) (Sheehan et al., 1998) for adults at baseline and the second assessment (Supplementary material), with 10 items each used to calculate sum scores (Table S1). For children, depression and anxiety were measured at baseline using the Child Behavior Checklist (Achenbach, 1991a) and the Youth Self-Report questionnaires (Achenbach, 1991b), with 13 items for depression and 6 for anxiety (Table S2). During the baseline visit, height, weight, waist and hip circumference of participants were measured (Sijtsma et al., 2021). BMI was calculated as weight (kg) divided by square of height (m^2^). Waist-hip-ratio (WHR) was calculated as waist circumference (cm) divided by hip circumference (cm). Number of cigarettes per day was measured at baseline by self-report questionnaire (Lifelines, 2022), and defined as the number of cigarettes of participants smoked each day. Daily alcohol intake (grams) was measured at baseline based on the food frequency questionnaire (Brouwer-Brolsma et al., 2021), and defined as average grams of alcohol drank by participants each day (Wang et al., 2022).

### Socio-economic status

Socio-economic status was measured at three levels: individual (educational attainment and occupational status), household (disposable household income), and neighborhood (NSES). Educational attainment was measured at baseline by self-report to the question “What is the highest level of education you have attained?” Occupational status was measured at baseline using the occupational prestige scale based on the International Standard Classification of Occupations (ISCO-08) (Ganzeboom & Treiman, 2003). Household income was measured at baseline by self-report to the question “what is the net income per month?” Disposable household income equals the average household income divided by equivalence factors based on the number of family members from the Statistics Netherlands website (Statistics-Netherlands, 2019). NSES was assessed by the Statistics Netherlands status score of the neighborhood of the home of the participant. Correlations observed between educational attainment, occupational status, and disposable household income were considerably larger (0.31-0.54, Table S3) than with NSES (0.08-0.09, Table S3). A confirmatory factor analysis of the three moderately correlated SES variables produced factor loadings of 0.73 for educational attainment, 0.74 for occupational status, and 0.42 for disposable household income. This latent SES factor was used in the primary analysis.

### Genetic data

Genome-wide genotyping was conducted on 55,063 participants, with 50,802 remaining after quality control. Following the exclusion of 34 individuals of non-European ancestry, the final sample was used for analysis. Details on quality control process and imputation are in the supplementary materials.

### Analytical approach

The aims of our analyses were twofold. First, to determine whether socio-economic status moderates genetic effects on depression, anxiety, obesity and substance use using linear mixed regression models to estimate main effects and interactions between PRSs and SES on individual outcomes. Second, to explore whether SES indices interact with shared genetic susceptibilities influencing the shared variance of these outcomes. To this end we used confirmatory factor analysis to estimate co-occurrence at the phenotypic level, and used genomic SEM to model the shared susceptibilities at the genetic level.

### Phenotypic and Genomic SEM

At the phenotypic level, confirmatory factor analysis was used to combine sum score of depression and anxiety to generate a latent factor representing the shared variance of depression and anxiety (DepAnx). Factor loadings were fixed to be equal for depression and anxiety. Similar analyses were conducted to model the shared variance between BMI and WHR (BmiWhr), and between smoking and alcohol use (SmkAlc). Finally, we combined DepAnx, BmiWhr and SmkAlc to generate one common higher order latent factor to represent the shared variance across all domains.

Genomic SEM models the genetic covariance structure of GWAS summary statistics using LD score regression to estimate the association of each SNP with the factor (a latent variable corresponding to the shared genetic variance between two GWASs) (Grotzinger et al., 2019). We used genomic SEM to combine depression and anxiety GWASs into a shared GWAS representing the shared genetic variance of co-occurring depression and anxiety (DepAnx). In genomic SEM, factor loadings were fixed to be equal between depression and anxiety GWAS. Similar shared GWASs were generated for co-occurring high BMI and WHR (BmiWhr), and smoking and alcohol use (SmkAlc). We then estimated a shared GWAS for a common factor of DepAnx, BmiWhr and SmkAlc. Figure S1 shows the genomic SEM path diagram. In genomic SEM, Q_SNP_ is a D^2^-distributed test statistic, which tests the heterogeneity of effect sizes of each SNP across each individual phenotype and the common factor (Grotzinger et al., 2019), providing insight into whether genetic aggregation beyond these domains is supported. Thus, the Q_SNP_ test statistic was used to examine the heterogeneity of SNP effects for DepAnx, BmiWhr, SmkAlc and the common higher order factor.

### Polygenic risk scores and statistical analysis

PRSs were generated by PLINK v1.9 (Purcell et al., 2007) and R 4.0.3 (R-Core-Team, 2013), and were calculated using the genotype data of the Lifelines participants and recent large summary statistics of GWAS meta-analysis for depression (Howard et al., 2019), anxiety (Purves et al., 2020), obesity (Pulit et al., 2019), and substance use (Liu et al., 2019) (Table S4), and shared GWASs for DepAnx, BmiWhr, SmkAlc and the single common factor. Strand-ambiguous SNPs were removed, and independent SNPs were obtained via clumping (r²=0.1, 1000 kb window). PRSs were calculated by weighting independent risk alleles by allelic effect sizes and aggregating them at eleven P thresholds (< 5×10^−8^, < 1×10^−7^, < 1×10^−6^, < 1×10^−5^, < 1×10^−4^, < 0.001, < 0.01, < 0.05, < 0.1, < 0.5, ≤ 1.0). Principal component analysis (PCA) was performed on the resulting PRSs, with the first PRS-PC used in all analyses (Coombes et al., 2020).

Linear mixed regression models were used to estimate interactions between SES and PRSs for individual outcomes, and interactions between SES and shared PRSs for co-occurrence of these outcomes with adjustment for family relationships in ASReml-R (Butler et al., 2018). Age, sex, chips (CytoSNP or GSA), and 10 principal components were covariates. Gene-environment correlations were tested using Pearson correlations between PRSs and SES, and phenotypic correlations and genetic correlations between outcomes were estimated using Pearson correlations and LD score regression. Most outcomes had skewed residuals, except for BMI, WHR, and BmiWhr (Figure S2). Sensitivity analyses were conducted by applying an inverse normal transformation to residuals to assess deviations from normality. The PRS by SES interactions were considered significant at *p* < 0.05. Attrition analyses compared demographic characteristics between participants with and without missing data.

## Results

Among all participants of Lifelines, 50,761 provided information on both genetic data and at least one phenotype (Figure S3). Table 1 shows their characteristics, with a mean age of 42.6 years, and 58.5% being female. Table S5 shows missing data ranging from 0.26% for BMI to 19.89% for the common factor. Males had more missing data on smoking, and younger participants had more missing data on obesity, while older participants had more missing data on educational attainment and occupational status. The findings in Table S6 demonstrate the outcomes of the attrition analyses conducted on a subset of participants without missing data. The results indicate that the interaction effects between PRSs (the shared PRS) and SES for both individual and aggregated outcomes were very similar to those obtained from the total sample.

**Table 1.** Characteristics of participants in the current study (n=50,761)

| Variables | n | mean±SD /median(IQR) |
| --- | --- | --- |
| Age | 50,761 | 42.63 ± 15.50 |
| Sex (female), n (%) | 50,761 | 29,673 (58.46%) |
| Outcome |  |  |
| Sum score of depression | 42,699 | 0.00 (0.00-1.00) |
| Sum score of anxiety | 42,588 | 0.00 (0.00-2.00) |
| Number of cigarettes per day | 48,603 | 0.00 (0.00-10.00) |
| Daily alcohol intake (grams) | 49,930 | 3.74 (0.69-10.39) |
| Body mass index (kg/m<sup>2</sup>) | 50,628 | 25.39 ± 4.51 |
| Waist-hip-ratio | 50,625 | 0.90 ± 0.08 |
| Comorbidity |  |  |
| DepAnx | 42,318 | 0.65 (0.00-2.32) |
| SmkAlc | 48,448 | 4.36 (0.80-9.60) |
| BmiWhr | 50,625 | 6.15 ± 0.62 |
| Common factor | 40,667 | 4.89 ± 0.81 |
| Socio-economic status |  |  |
| Educational attainment (years) | 50,204 | 14.22 ± 4.12 |
| Disposable household income (euros/month) | 45,214 | 1611.98 ± 514.40 |
| Occupational status | 48,914 | 43.73 ± 13.03 |
| Neighborhood socio-economic status | 49,381 | -0.53 ± 1.05 |
| Latent SES factor | 43,463 | 0.00 ± 1.48 |

Abbreviations: SD, standard deviation; IQR, interquartile range. DepAnx, the co-occurrence of depression and anxiety; SmkAlc, the co-occurrence of number of cigarettes per day and daily alcohol intake; BmiWhr, the co-occurrence of high BMI and WHR; Common factor, the co-occurrence of DepAnx, SmkAlc, and BmiWhr. Latent SES factor: a latent SES factor combining educational attainment, occupational status, and disposable household income in confirmatory factor analysis.

Polygenic risk scores for individual outcomes explained 0.74%, 0.80%, 1.04%, 2.19%, 2.36%, and 8.63% for anxiety, depression, daily alcohol intake, WHR, number of cigarettes per day, and BMI, respectively (Figure S4-7). The shared PRSs explained 0.62% for DepAnx, 5.36% for BmiWhr, 1.85% for SmkAlc, and 3.29% for the higher order common factor (Figure S3-6). Figure S8 and Table S7 shows the variance explained by 4 individual SES indices and one latent SES factor. For DepAnx and SmkAlc domains, SES explained less variance at the aggregated level than for the variance of at least one individual outcome. For the BmiWhr domain, SES explained more variance at the aggregated than at the specific BMI and WHR levels. Educational attainment, occupational status, disposable household income, NSES, and the latent SES factor explained 1.94%, 1.15%, 0.75%, 0.25%, 2.16% of the higher order common factor variance (Figure S8 and Table S7).

Lower socio-economic status amplified the genetic effects on depression, anxiety, BMI, and smoking, while no interactions between the latent SES factor and PRSs were found for WHR (*p*=0.07) and alcohol use (*p*=0.67) (Figure 1-3 and Table S7). In terms of variance explained, the main effects of PRSs and the latent SES factor, along with their interactions, accounted for varying proportions of variance across individual level outcomes: 2.44% for depression, 1.62% for anxiety, 9.47% for BMI, 2.63% for WHR, 3.84% for smoking, and 1.20% for alcohol use (Table S7). In further analysis of individual, household, and neighborhood level SES indices, fourteen of twenty-four PRSs-by-SES interactions showed significant results (*ps*<0.05) for depression, anxiety, BMI, smoking, and alcohol use, but not for WHR (Figure S9-11 and Table S6). However, higher disposable household income amplified the PRS effect on alcohol use (*p*=0.04). At the neighborhood level, lower NSES amplified the genetic effects on depression and BMI (*ps*<0.05), while no interactions were detected between NSES and PRSs for anxiety, WHR, smoking and alcohol use.

**Figure 1.**
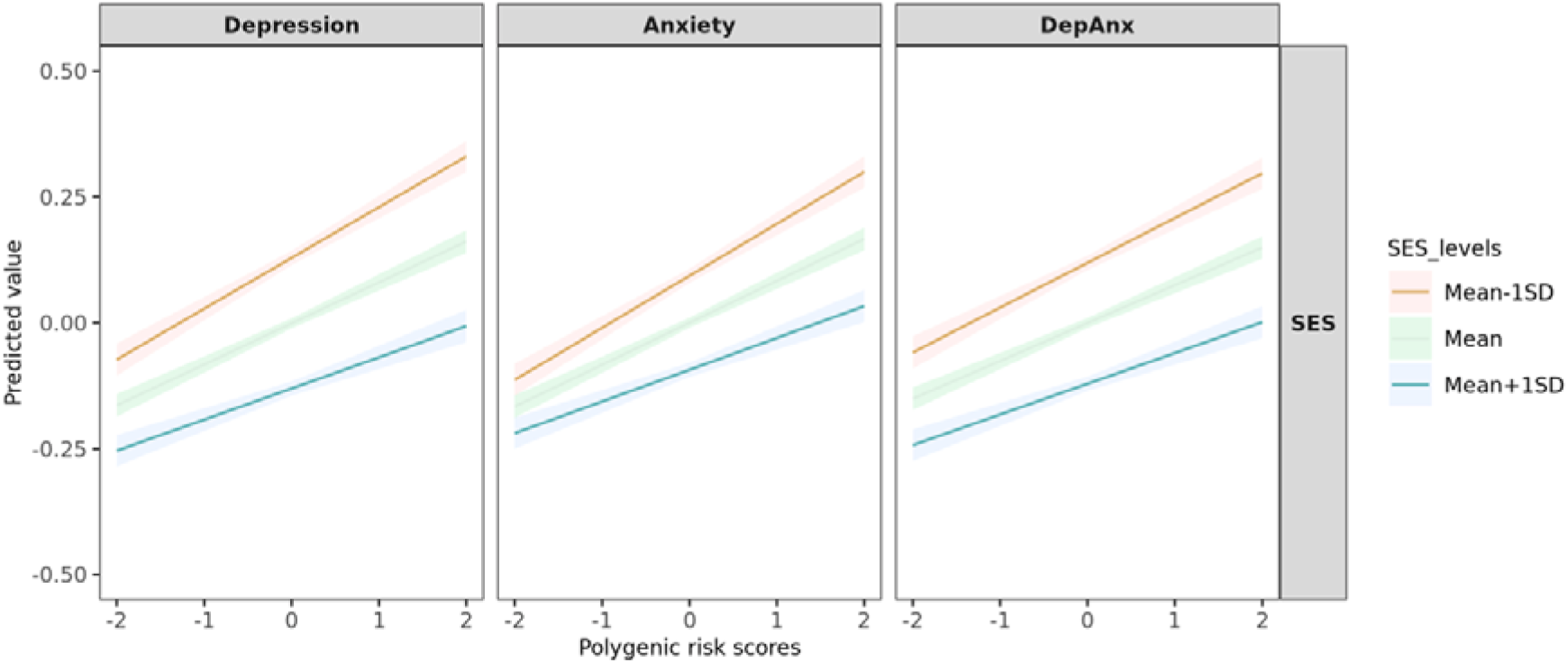
The interactions between PRSs and SES for depression, anxiety and DepAnx. Abbreviations: DepAnx, the co-occurrence of depression and anxiety. Y axis represents the standardized scores of depression, anxiety and DepAnx. The linear mixed regression model was adjusted for age, sex, chip (CytoSNP or GSA) and 10 principal components.

**Figure 2.**
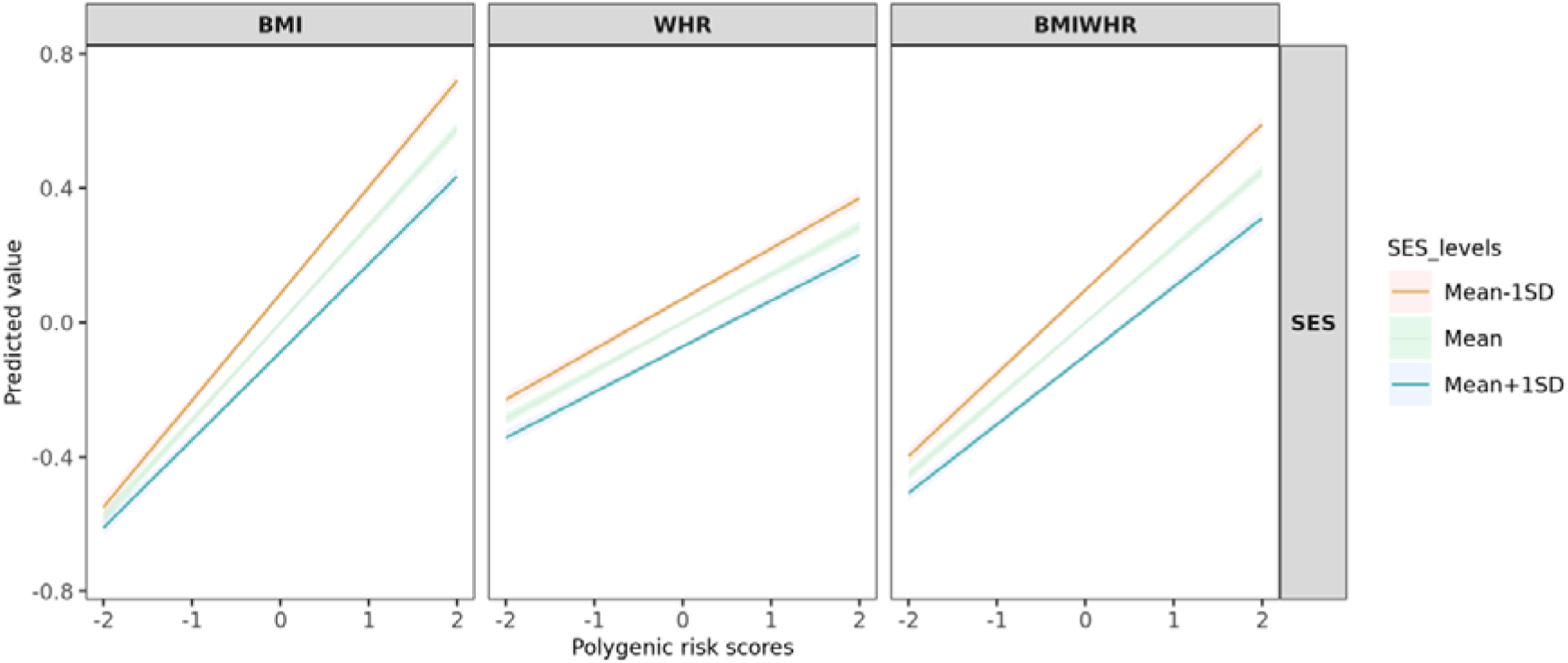
The interactions between PRSs and SES for BMI, WHR and BmiWhr. Abbreviations: BMIWHR, the co-occurrence of BMI and WHR. Y axis represents the standardized scores of BMI, WHR and BMIWHR. The linear mixed regression model was adjusted for age, sex, chip (CytoSNP or GSA) and 10 principal components.

**Figure 3.**
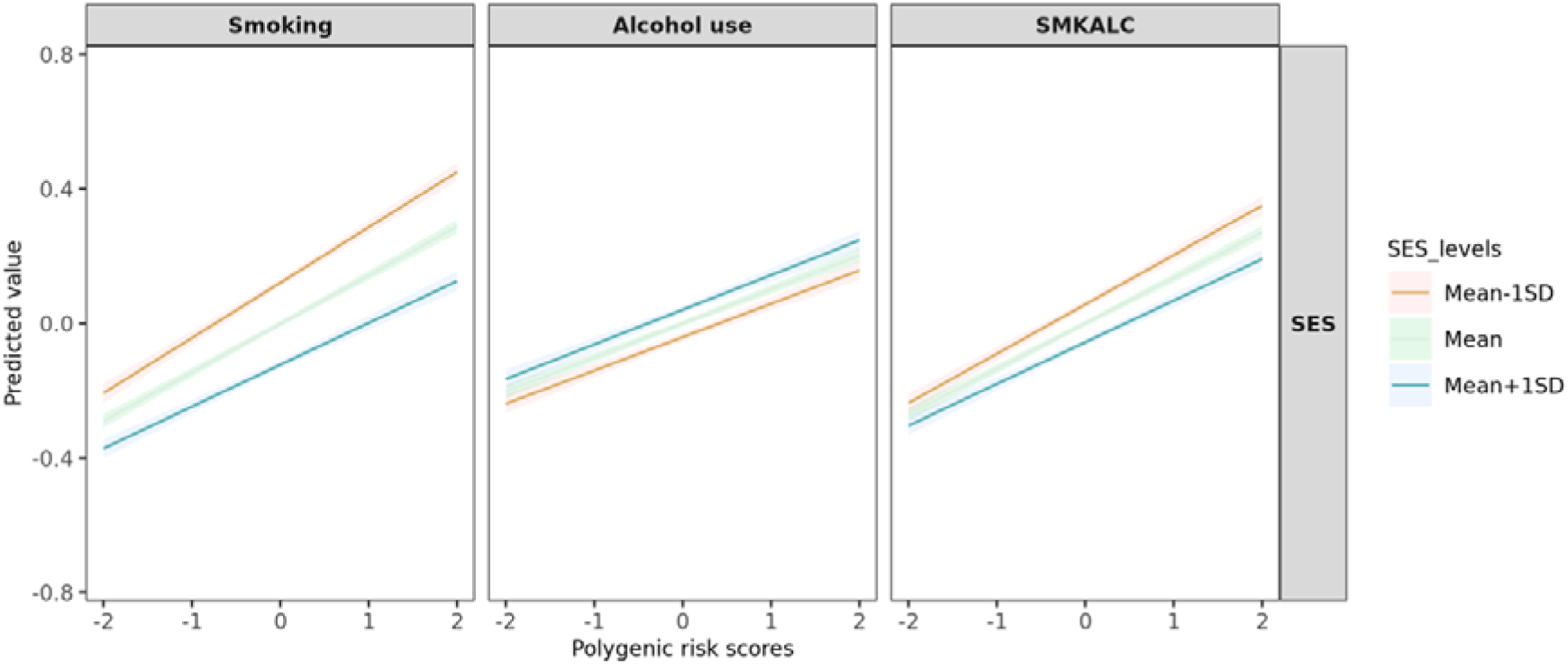
The interactions between PRSs and SES for alcohol use, smoking and SmkAlc. Abbreviations: SMKALC, the co-occurrence of smoking and alcohol use. Y axis represents the standardized scores of alcohol use, smoking and SMKALC. Linear mixed regression model adjusted for age, sex, chip (CytoSNP or GSA) and 10 principal components.

At the aggregated outcome level, lower SES amplified the shared PRSs effects on DepAnx, BmiWhr, SmkAlc, as well as the common factor (*ps*<0.05) (Figure 1-4 and Table S7). However, the interaction effect sizes tended to be smaller than those for individual outcomes. Regarding the variance explained, the main effects of shared PRSs and the latent SES factor, along with their interactions, accounted for varying proportions of variance across aggregated level outcomes: 2.04% for DepAnx, 6.35% for BmiWhr, 2.18% for SmkAlc, and 5.09% for the common factor (Table S7). Further analysis of individual, household, and neighborhood-level SES indices revealed that nine out of sixteen interactions between SES and shared PRSs were significant (*ps*<0.05).

**Figure 4.**
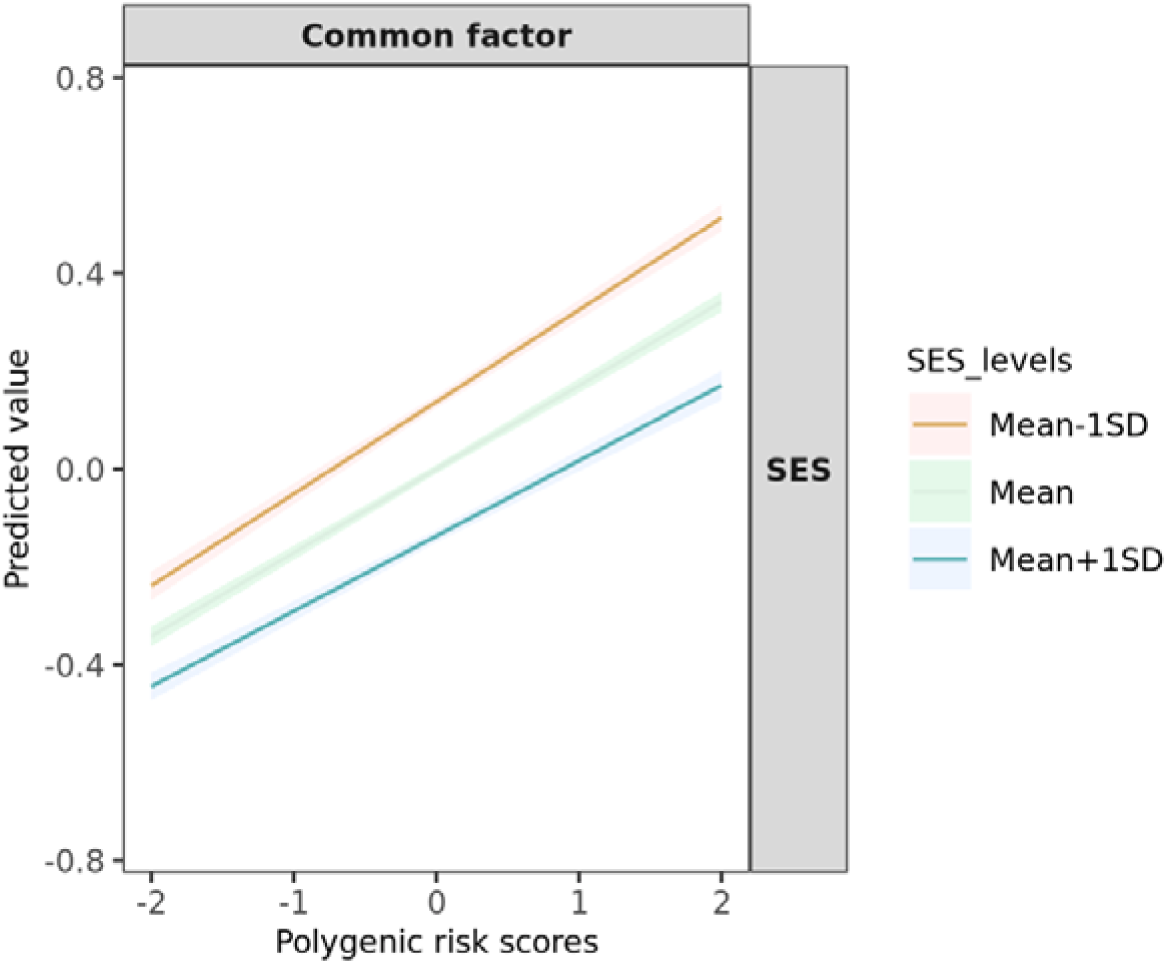
The interactions between PRSs and SES for the common factor. Abbreviations: Common factor, the co-occurrence of depression, anxiety, obesity and substance use. Y axis represents the standardized scores of the common factor. The linear mixed regression model was adjusted for age, sex, chip (CytoSNP or GSA) and 10 principal components.

To assess the impact of the skewed distribution of outcomes on interactions, Table S8 shows that after applying the inverse normal transformation of the covariate residualized outcomes, the interaction effects between PRSs and SES for the individual outcomes were similar but generally less significant. Further, the interaction effect between the shared PRS and SES on BmiWhr became more significant, but the interactions became non-significant for DepAnx, SmkAlc and the common factor. These findings suggest that the distribution of outcomes had a minor effect on the interaction effects between PRSs and SES for individual outcomes, but a more pronounced effect on the interaction effects between the shared PRSs and SES for the co-occurrence of outcomes. Table S9 shows the Pearson correlations between PRSs (shared PRSs) and SES were small but significant (ranging from -0.09 to 0.02), indicating a small gene-environment correlation between PRSs and SES. Table S10 presents the genetic and phenotypic correlations between outcomes. We found moderate to high genetic correlations, ranging from 0.14 between BmiWhr and SmkAlc to 0.90 between depression and anxiety. Additionally, we observed small to moderate phenotypic correlations, ranging from 0.02 between DepAnx and BmiWhr to 0.67 between depression and anxiety. Table S11 displays the number of SNPs with heterogeneity at various thresholds in the QSNP test for DepAnx, BmiWhr, SmkAlc, and the common higher order factor. Only a small proportion of SNPs (0.06% or 3,340 SNPs) were identified as high heterogeneity SNPs at the QSNP P-value<5×10^−8^ threshold. When comparing the variance explained by the shared PRS including all SNPs (3.29%) to the variance explained by the shared PRS only including SNPs with low heterogeneity (QSNP p>5×10^−3^), a similar amount of variance explained was observed for the common factor (3.28%). These results indicate that the effect of excluding heterogeneous SNPs from the shared PRS is minor. (Figure S13).

## Discussion

In this large and comprehensive study on polygenic risk-by-SES interaction effects, we found that lower socio-economic status amplified genetic susceptibilities for individual and aggregated outcomes of depression, anxiety, BMI, WHR, smoking, and alcohol use. Eight of ten PRSs-by-latent SES factor interactions were significant (*ps*<0.05) for depression, anxiety, BMI, smoking, and aggregated outcome level, but not for WHR (*p*=0.07) and alcohol use (*p*=0.67). At the aggregated outcome level, interaction effects were mostly smaller than effects for individual outcomes.

Our study found that lower SES amplified the genetic effects on depression, anxiety, BMI and smoking, consistent with previous findings (Agerbo et al., 2021, Barcellos et al., 2018, Pasman et al., 2020, Thompson et al., 2020). Unlike prior studies that usually focused on one specific SES index [e.g. educational attainment in relation to obesity (Barcellos et al., 2018)], our results confirmed significant polygenic risk-by-SES interactions at individual, household, and neighborhood levels for depression and BMI. We observed significant PRS-by-SES interactions for BMI across four SES indices. Barcellos et al. found a significant interaction effect between PRS for BMI and educational attainment on obesity, but not on BMI (n=253,715) (Barcellos et al., 2018), which might be attributed to the previous underpowered GIANT 2015 GWAS (236,781 Europeans) (Locke et al., 2015) compared to the more recent larger GWAS used in the present study (806,834 Europeans) (Pulit et al., 2019). Our study also found that lower educational attainment and lower occupational status significantly interacted with PRS for smoking, but not for disposable household income and NSES. Likewise, Pasman et al found no interaction between PRS and SES (housing value and monthly income) for smoking in a Dutch study (n=6,471) (Pasman et al., 2020). Higher disposable household income slightly amplified the genetic effect on alcohol use (*p*=0.04), in line with a previous finding that higher SES (housing value and monthly income) amplified the PRS effect on alcohol use (*p*=0.005) (Pasman et al., 2020). Both studies suggest that genetic risk may exacerbate alcohol use in higher income individuals, though lower SES is associated with more severe alcohol-related consequences (e.g., incident alcohol use disorder and mortality (Collins, 2016, Lasserre et al., 2022).

Our study is the first to explore polygenic risk-by-SES interaction at the aggregated phenotypic level. We found that lower SES amplifies shared genetic susceptibility across disease domains, although only one of twenty interaction effects (four for the latent SES factors, twelve for the three domains and four for the common factor across 4 SES indices) showed slightly stronger effects than those observed for individual outcomes. The exception was that lower NSES had a stronger interaction with the PRS derived from the shared genetic part between depression and anxiety than with the individual PRSs (Figure S9). Additionally, shared PRSs explained less variance in aggregated outcomes (e.g. 0.62% for DepAnx) than in individual outcomes (e.g. 0.74% for anxiety and 0.80% for depression, Figure S4). In line with this, a recent study applying genomic SEM across 11 psychiatric disorders found a single dimension of genetic risk cannot represent 11 different psychiatric disorders (Grotzinger et al., 2022). The finding suggested that (part of the) shared pleiotropic loci (or SNPs) may act heterogeneously across different individual outcomes, which may produce weaker joint effects of these loci on aggregated outcomes. However, in the present study, the Q_SNP_ test for the common factor showed minimal SNPs heterogeneity across domains (less than 0.1%, Q_SNP_ p<5×10^−8^). A PRS based on SNPs with low heterogeneity (Q_SNP_ p>5×10^−3^) explained a similar amount of variance for the common factor as a PRS including all SNPs (3.28% vs 3.29%, Figure S13). These findings indicate that SNP heterogeneity does not fully account for the weaker main and interaction effects with SES at the aggregated level. Another reason for weaker effects at the aggregated level is that SES has inverse effects on smoking and alcohol use - lower SES is linked to heavier smoking, while higher SES is linked to heavier alcohol use-attenuating effects when these are combined. Despite psychiatric and somatic diseases often co-occurring and sharing genetic background, our findings suggest that studying G×E interactions at an aggregated disease level does not enhances etiological understanding of frequently co-occurring issues.

SES is a complex and multifactorial construct, so it is unclear what exactly the mechanisms are of SES moderating the genetic susceptibility on health outcomes. While our study focused on SES as a risk factor for poor mental and physical health, it is worth noting that poor health itself may also lead to lower SES, such as through work disability (Nieuwenhuijsen et al., 2020). Interpreting polygenic risk-by-SES effects is further complicated by the genetic component of SES. For example, PRS explain 12%-16% of the variance in educational attainment (Okbay et al., 2022), and genetic correlations exist between SES and psychiatric outcomes (rG for SES was -0.52 with anxiety, -0.42 with major depression, -0.25 with alcohol use quantity, and -0.50 with smoking initiation) (Marees et al., 2021). Gene-environment correlation (rGE) effects (Jaffee & Price, 2007), wherein an individual’s genetic make-up may shape their environmental exposures, potentially confounds gene-environment interaction findings (Pasman et al., 2020). Recent studies suggest using family designs to quantify rGE effects (Kong et al., 2018, Selzam et al., 2019), also known as genetic nurture. For instance, assessing the impact of non-transmitted parental alleles on offspring health can reveal the influence of parental rearing environments (Kong et al., 2018). Distinguishing between within- and between-family PRS effects among siblings can also help quantify rGE effects, with SES playing a significant role in between family component with cognitive traits, such as educational attainment and intelligence, but not with non-cognitive traits, such as BMI and body height (Selzam et al., 2019). Although we could not control for the rGE in this study, correlations between PRSs and SES indices, although significant, were small (Table S9), indicating minimal bias in the G×E interaction effects. A simulation study by Akimova and colleagues (Akimova et al., 2021) supports this, showing slight attenuation of main effects but less for interaction effects. Advanced methods such as genomic SEM (Grotzinger et al., 2019), can remove the genetic variance of environmental exposures such as SES from PRSs, helping future studies better estimate interactions between environmental exposures and PRSs while controlling for the genetic influence on environmental exposures. For example, interaction between PRS and neighborhood income for lifetime smoking became non-significant when SNP-based genetic educational attainment variance was removed (Pasman et al., 2022), indicating the need for further research.

Our study has several limitations. First, the depression score was mainly based on symptoms from the past 2 weeks and anxiety from the past 6 months, likely underestimating the effects of polygenic risk-by-SES interactions compared to lifetime outcomes. Second, G×E interactions may be scale-dependent (Coleman et al., 2020). However, sensitivity analyses using inverse normal transformation confirmed the robustness of the findings and showed that the interactions were still significant for individual outcomes, though not for their co-occurrence (except for BmiWhr, Table S8). Third, we did not control for rGE, which may have confounded our G×E interaction findings. However, we found gene-environment correlations of relatively minor magnitude, indicating that any bias caused by rGE on the G×E interaction effects is likely to have been small. Fourth, our study was conducted only among Europeans, which make it difficult to generalize to other ancestries.

Based on large sample size and a variety of SES indices, our study provides evidence of the presence of polygenic risk-by-SES interactions at the level of individual PRSs for depression, anxiety, BMI, smoking, and alcohol use. However, smaller shared PRS and smaller PRS×SES effects within and across disease domains compared to individual outcomes suggest limited utility in understanding of shared etiological factors. It is noteworthy that effect sizes of the interaction between SES and PRS remain relatively modest, indicating that the clinical applicability of our findings is currently limited. Notably, for obesity, the current PRS explains about 9% of the variance in BMI, suggesting that interventions targeting individuals with high genetic risk to obesity and low SES backgrounds may soon be feasible. Strategies such as promoting healthy dietary habits and increasing physical activity could then be tailored to prevent the onset of obesity in high-risk groups. Advances in GWAS and PRS methodologies (e.g., MegaPRS (Zhang et al., 2021) and SBayesRC (Zheng et al., 2024)) may reveal stronger G×E interaction effects may be found, which may support targeted prevention for populations that have both high genetic risk and low socio-economic status. However, ethical concerns, such as fear or misunderstanding upon returning genetic information and the risk of (self-) stigma and scapegoating (Baker et al., 2022, Baty & Davis, 2023, Hallquist et al., 2021, Weitzman et al., 2024), mush be considered when targeting specific high-risk groups, balance the benefits of tailored prevention against all ethical complications.

## Data Availability

All data produced in the present work are contained in the manuscript

https://wiki.lifelines.nl/doku.php

## ACKNOWLEDGEMENTS

We acknowledge the services of the Lifelines Cohort Study, the contributing research centres delivering data to Lifelines, and all the study participants.

## AUTHOR CONTRIBUTIONS

RW, HS, and CAH contributed to the study conception and design. Lifelines Cohort Study offered the data. RW did data analysis and drafted the manuscript. All authors participated in revising it critically for important intellectual content.

