## Supplementary for "Polygenic risk-by-socio-economic status interaction effects on specific and aggregated outcomes of depression, anxiety, body mass index, waist-hip-ratio, smoking and alcohol use"

^2^A full list of members and affiliations appears in the Supplementary Note

^3^Department of Psychiatry, University of Groningen, University Medical Center Groningen, Groningen, Netherlands

**Supplementary**

**Supplementary Methods**

**Measurements**

Current depression and anxiety were measured using the MINI International Neuropsychiatric Interview (MINI) [1] for adults at baseline and the second assessment. The MINI was performed as an individual face-to-face interview by a trained research nurse at baseline when participants visited a Lifelines research facility. During the follow-up, the MINI was administered as a digital questionnaire with participants entering their answers under the supervision of a trained research nurse on location. In the early stages of the baseline measurement wave, “skips” were used in the MINI interview such that some questions were asked, or not asked, depending on the participants’ responses on screening questions. In order to collect full data on all participants, skips were removed from the MINI at a later stage of the baseline measurement. To capture anxiety and depression as a continuous outcome, we used the MINI without skips at follow-up for participants who had been assessed using the MINI with skips at baseline. The depression sum score captured current major depression in the past two weeks, measured by nine items, and one item for dysthymia in the past two years. The anxiety sum score encompassed four types of anxiety, with a focus on generalized anxiety disorder in the past six months, measured by seven items. Additionally, there was one item each for panic disorder, agoraphobia, and social anxiety disorder in the past month.

**Table S1. Items for depression and anxiety from the MINI**

| Phenotypes | Items | Questions | Answers |
| --- | --- | --- | --- |
| Sum score of depression for adults | A1 | Have you been consistently depressed or down, most of the day, nearly every day, for the past two weeks? | 0=no  1=yes |
|  | A2 | In the past two weeks, have you been much less interested in most things or much less able to enjoy the things you used to enjoy most of the time? |  |
|  | A3A | Was your appetite decreased or increased nearly every day? Did your weight decrease or increase without trying intentionally? |  |
|  | A3B | Did you have trouble sleeping nearly every night (difficulty falling asleep, waking up in the middle of the night, early morning wakening or sleeping excessively)? |  |
|  | A3C | Did you talk or move more slowly than normal or were you fidgety, restless or having trouble sitting still almost every day? |  |
|  | A3D | Did you feel tired or without energy almost every day? |  |
|  | A3E | Did you feel worthless or guilty almost every day? |  |
|  | A3F | Did you have difficulty concentrating or making decisions almost every day? |  |
|  | A3G | Did you repeatedly consider hurting yourself, feel suicidal, or wish that you were dead? |  |
|  | B1 | Have you felt sad, low or depressed most of the time for the last two years? |  |
| Sum score of anxiety for adults | O1A | Have you worried excessively or been anxious about several problems of daily life (problems at work, at home or in your close circle) over the past 6 months? | 0=no  1=yes |
|  | O3A | When you were anxious over the past 6 months, did you, most of the time, feel restless, keyed up or on edge? |  |
|  | O3B | When you were anxious over the past 6 months, did you, most of the time, feel tense? |  |
|  | O3C | When you were anxious over the past 6 months, did you, most of the time, feel tired, weak or exhausted easily? |  |
|  | O3D | When you were anxious over the past 6 months, did you, most of the time, have difficulty concentrating or find your mind going blank? |  |
|  | O3E | When you were anxious over the past 6 months, did you, most of the time, feel irritable? |  |
|  | O3F | When you were anxious over the past 6 months, did you, most of the time, have difficulty sleeping (difficulty falling asleep, waking up in the middle of the night, early morning wakening or sleeping excessively)? |  |
|  | E1 | Have you, on more than one occasion, had spells or attacks when you suddenly felt anxious, frightened, uncomfortable or uneasy, even in situations where most people would not feel that way? |  |
|  | F1 | Do you feel anxious or uneasy in places or situations where you might have a panic attack or the panic-like symptoms we just spoke about, or where help might not be available or escape might be difficult: like being in a crowd, standing in a line (queue), when you are alone away from home or alone at home, or when crossing a bridge, traveling in a bus, train or car? |  |
|  | G1 | In the past month, were you fearful or embarrassed being watched, being the focus of attention, or fearful of being humiliated? This includes things like speaking in public, eating in public or with others, writing while someone watches, or being in social situations. |  |

For children, depression (n=2,864) and anxiety (n=2,865) were measured using children’s behavior questionnaires at baseline, combining the Child Behavior Checklist (CBCL) [2] for ages 8-17 years and the Youth Self-Report (YSR) [3] for 13-17 years. Sum scores of depression and anxiety were calculated as continuous traits (Items are in the Table S2).

**Table S2. Items for calculating sum scores of depression and anxiety in CBCL and YSR**

| Phenotypes | Age | Items | Questions | Answers |
| --- | --- | --- | --- | --- |
| Sum score of depression for children | Parent-report CBCL  (8-17 years) | CBCL5 | There are not many things it likes | 0=not at all  1=a little bit or sometimes  2=clearly or often |
|  |  | CBCL14 | Cries a lot |  |
|  |  | CBCL18 | Self-harms, or tries to commit suicide |  |
|  |  | CBCL24 | Does not eat well |  |
|  |  | CBCL35 | Feels useless or inferior |  |
|  |  | CBCL52 | Feels very guilty |  |
|  |  | CBCL54 | Is very tired without reason |  |
|  |  | CBCL76 | Sleeps less than most boys and girls |  |
|  |  | CBCL77 | Sleeps more than most boys and girls |  |
|  |  | CBCL91 | Talks about wanting to kill itself |  |
|  |  | CBCL100 | Problems sleeping |  |
|  |  | CBCL102 | Is not very active, moves slowly or has little energy |  |
|  |  | CBCL103 | Is unhappy, sad or depressed |  |
|  | Self-report YSR  (13-17 years) | ACHYSR5 | There is little that I like |  |
|  |  | ACHYSR14 | I cry a lot |  |
|  |  | ACHYSR18 | I try to deliberately hurt or kill myself |  |
|  |  | ACHYSR24 | I do not eat as well as I should |  |
|  |  | ACHYSR35 | I feel useless or inferior |  |
|  |  | ACHYSR52 | I feel very guilty |  |
|  |  | ACHYSR54 | I feel very tired without knowing why |  |
|  |  | ACHYSR83 | I sleep less than most boys and girls |  |
|  |  | ACHYSR84 | I sleep more than most boys and girls |  |
|  |  | ACHYSR98 | I think about killing myself |  |
|  |  | ACHYSR107 | I have trouble sleeping |  |
|  |  | ACHYSR109 | I don't have a lot of energy |  |
|  |  | ACHYSR110 | I am unhappy, sad or depressed |  |
| Sum score of anxiety for children | Parent-report CBCL  (8-17 years) | CBCL11 | Clings to adults or is too dependent | 0=not at all  1=a little bit or sometimes  2=clearly or often. |
|  |  | CBCL29 | Is afraid of certain animals, situations or locations other than school |  |
|  |  | CBCL30 | Is afraid to go to school |  |
|  |  | CBCL45 | Is nervous, twitchy or tense |  |
|  |  | CBCL50 | Is overly scared or anxious |  |
|  |  | CBCL112 | Worries |  |
|  | Self-report YSR  (13-17 years) | ACHYSR11 | I am too dependent on adults |  |
|  |  | ACHYSR29 | I am afraid of certain animals, situations or locations other than school |  |
|  |  | ACHYSR30 | I am afraid to go to school |  |
|  |  | ACHYSR45 | I am nervous, highly-strung or tense |  |
|  |  | ACHYSR50 | I am too frightened or scared |  |
|  |  | ACHYSR119 | I worry a lot |  |

**SES latent factor**

Given the moderate correlations observed between educational attainment, occupational status, and disposable household income (0.31-0.54, Table S3), whereas the correlations between NSES and other SES variables are relatively low (0.08-0.09, Table S3), we conducted a confirmatory factor analysis on the three SES variables with moderate correlations. Factor loadings were determined to be 0.73 for educational attainment, 0.74 for occupational status, and 0.42 for disposable household income. Subsequently, this derived latent SES factor was used in our primary analysis.

**Table S3. Spearman correlations between individual SES variables**

| Trait 1 | Trait 2 | Correlations | *P* |
| --- | --- | --- | --- |
| Education years | Occupational status | 0.54 | < 2.20×10^-16^ |
| Education years | Disposable household income | 0.31 | < 2.20×10^-16^ |
| Education years | Neighbourhood SES | 0.09 | < 2.20×10^-16^ |
| Occupational status | Disposable household income | 0.32 | < 2.20×10^-16^ |
| Occupational status | Neighbourhood SES | 0.09 | < 2.20×10^-16^ |
| Neighbourhood SES | Disposable household income | 0.08 | < 2.20×10^-16^ |

**Genetic data**

A total of 55,063 participants were selected for genome-wide genotyping. A first subset of 17,033 participants was genotyped using the Illumina CytoSNP-12v2 array [4]. Pre-imputation quality control was performed in which samples and variants were excluded with a call rate<95%, as well as variants with Hardy-Weinberg equilibrium (HWE) P<1×10-4, or minor allele frequency (MAF) <1%, and samples with a sex mismatch, deviating heterozygosity (>4 SD from the mean) or of non-European ancestry. A total of 15,400 samples and 265,000 single nucleotide polymorphisms (SNPs) were available for analysis. A second subset of 38,030 participants were genotyped using the Infinium Global Screening Array® (GSA) MultiEthnic Disease Version [4]. Standard quality control was performed on both samples and markers, including removal of samples and variants with a low genotyping call rate (<99%), variants showing deviation from HWE (P<1×10-6) or excess of Mendelian errors in families (>1% of the parent-offspring pairs), and samples with a sex mismatch, and very high or low heterozygosity. After quality control, a total of 36,339 samples and 571,420 SNPs were available for analysis. These two genotyping datasets were imputed using the HRC panel v1.1 at the Sanger imputation server [5], and variants with an imputation quality score higher than 0.4 for variants with a MAF >0.01 were retained. After removing duplicate samples between the two genetic datasets (n=937), 50,802 participants with genetic data were available. Among them, 34 were found to be non-European ancestry, and thus were excluded from our main analysis.

**GWASs and genomic SEM**

As shown in Figure S1, we combined the summary statistic from recent large GWAS (Table S4) to run genomic SEM at two levels. As data from some of the Lifelines subjects (n=8,118) was used in the GIANT 2015 GWAS meta-analysis for BMI and WHR [6], we used the MetaSubtract R package [7] to exclude the effect of Lifelines participants.

**Table S4. Summary statistics of recent large GWAS used in the present study**

| **Outcome** | **GWAS** | **Year** | **Datasets** | **Sample size** | **Phenotype** |
| --- | --- | --- | --- | --- | --- |
| Depression | Howard et al. [8] | 2019 | UKB+PGC | 500,199 | Lifetime depression |
| Anxiety | Purves et al. [9] | 2020 | UKB+iPSYCH+ANGST | 114,019 | Lifetime anxiety |
| BMI | Pulit et al. [10] | 2019 | UKB+GIANT-Lifelines | 798,716 | BMI |
| WHR | Pulit et al. [10] | 2019 | UKB+GIANT-Lifelines | 689,617 | WHR |
| Smoking | Liu et al. [11] | 2019 | GSCAN-23andMe | 632,802 | Ever smoking |
| Alcohol use | Liu et al. [11] | 2019 | GSCAN-23andMe | 537,349 | Drinks per week |

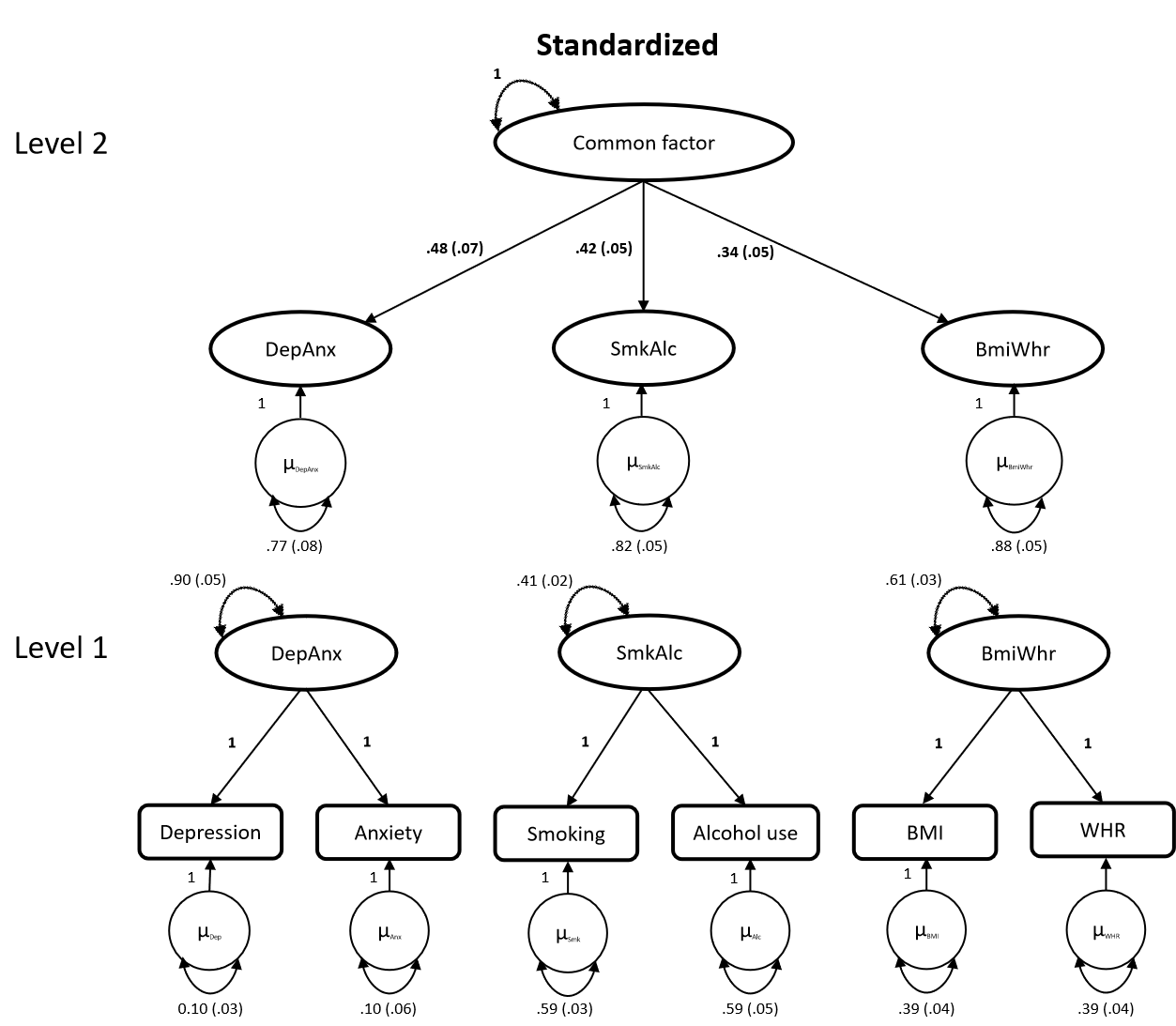

**Figure S1. Path diagram of Genomic SEM** Abbreviations: DepAnx, the co-occurrence of depression and anxiety; SmkAlc, the co-occurrence of number of cigarettes per day and daily alcohol intake; BmiWhr, the co-occurrence of BMI and WHR; Common factor, the co-occurrence of DepAnx, SmkAlc, and BmiWhr.

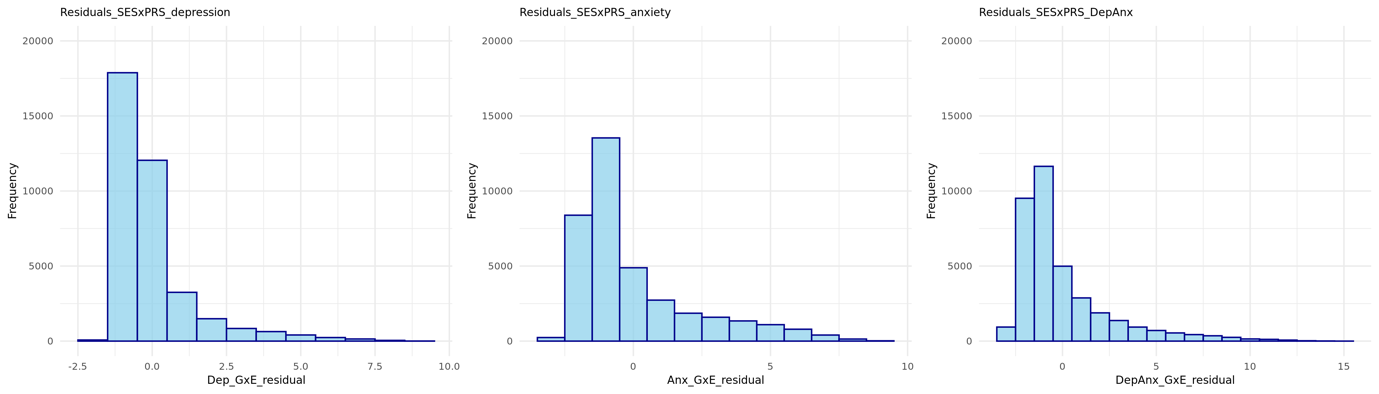

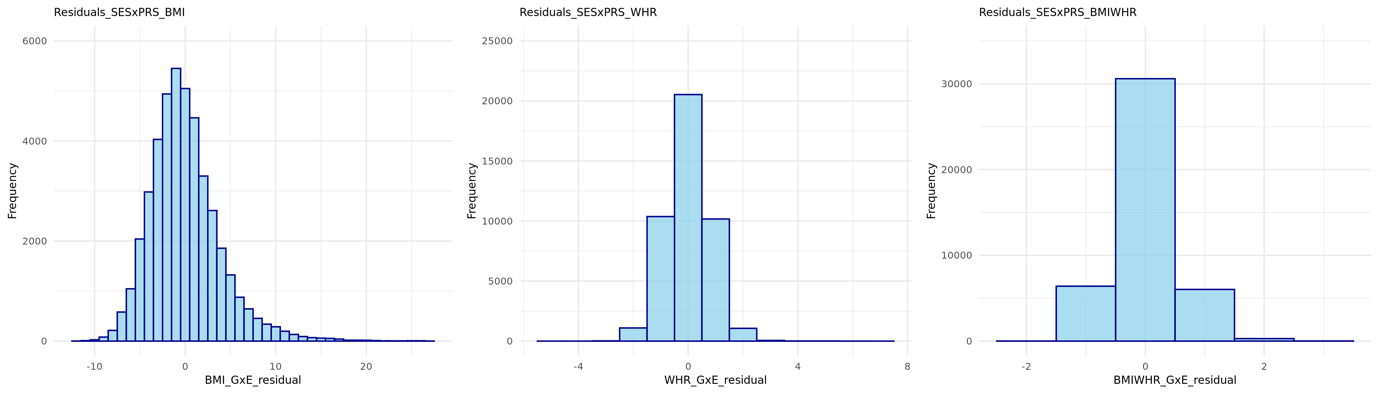

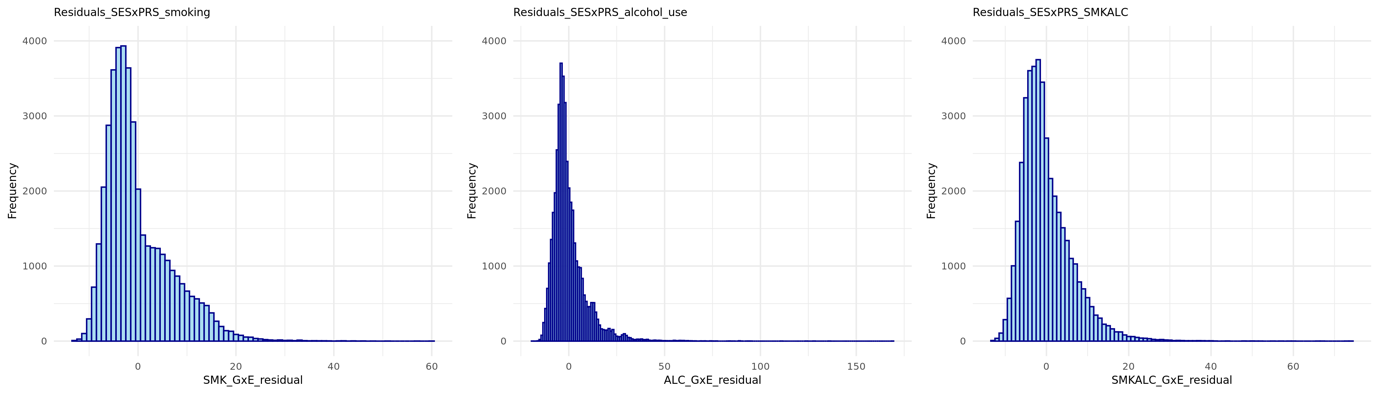

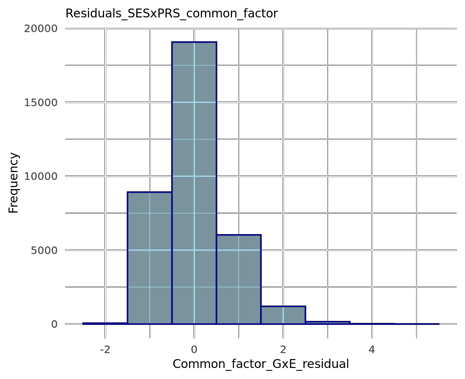

**Figure S2. The histogram of residuals of the models tested using linear regression**

Residuals are those of the full GxE models adjusted for SES, PRS, and SESxPRS, age, sex, chip type (CytoSNP or GSA), and 10 principal components.

**Without measurement of depression, anxiety, obesity, or substance use or non-European (n=41)**

**Phenotypic data**

**Genetic data in Lifelines**

**Final sample size**

**(n=50,761)**

**Genetic data**

**(n=50,802)**

**Overlap (n=937)**

**GSA QCed**

**(n=36,339)**

**CytoSNP QCed**

**(n=15,400)**

**GSA**

**(n=38,030)**

**CytoSNP**

**(n=17,033)**

**Figure S3. Flow chart** Abbreviations: CytoSNP, Illumina CytoSNP-12v2 array; GSA, Infinium Global Screening Array® (GSA) MultiEthnic Disease Version.

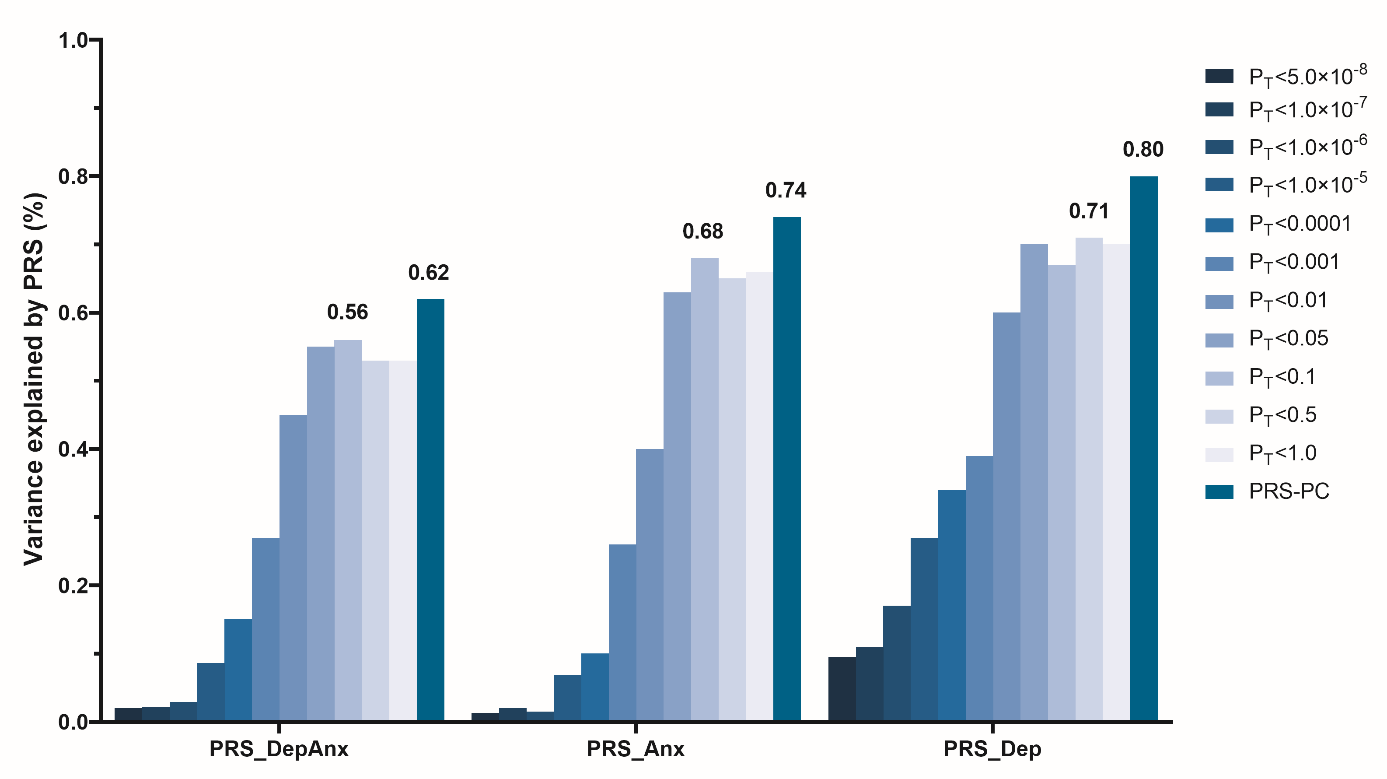

**Figure S4. Variance explained by PRSs for DepAnx, anxiety and depression** Abbreviations: DepAnx, the co-occurrence of depression and anxiety; PRS_DepAnx, the shared polygenic risk score for depression and anxiety; PRS_Dep, polygenic risk score for depression; PRS_Anx, polygenic risk score for anxiety; PRS-PC, the first principal component of PRSs.

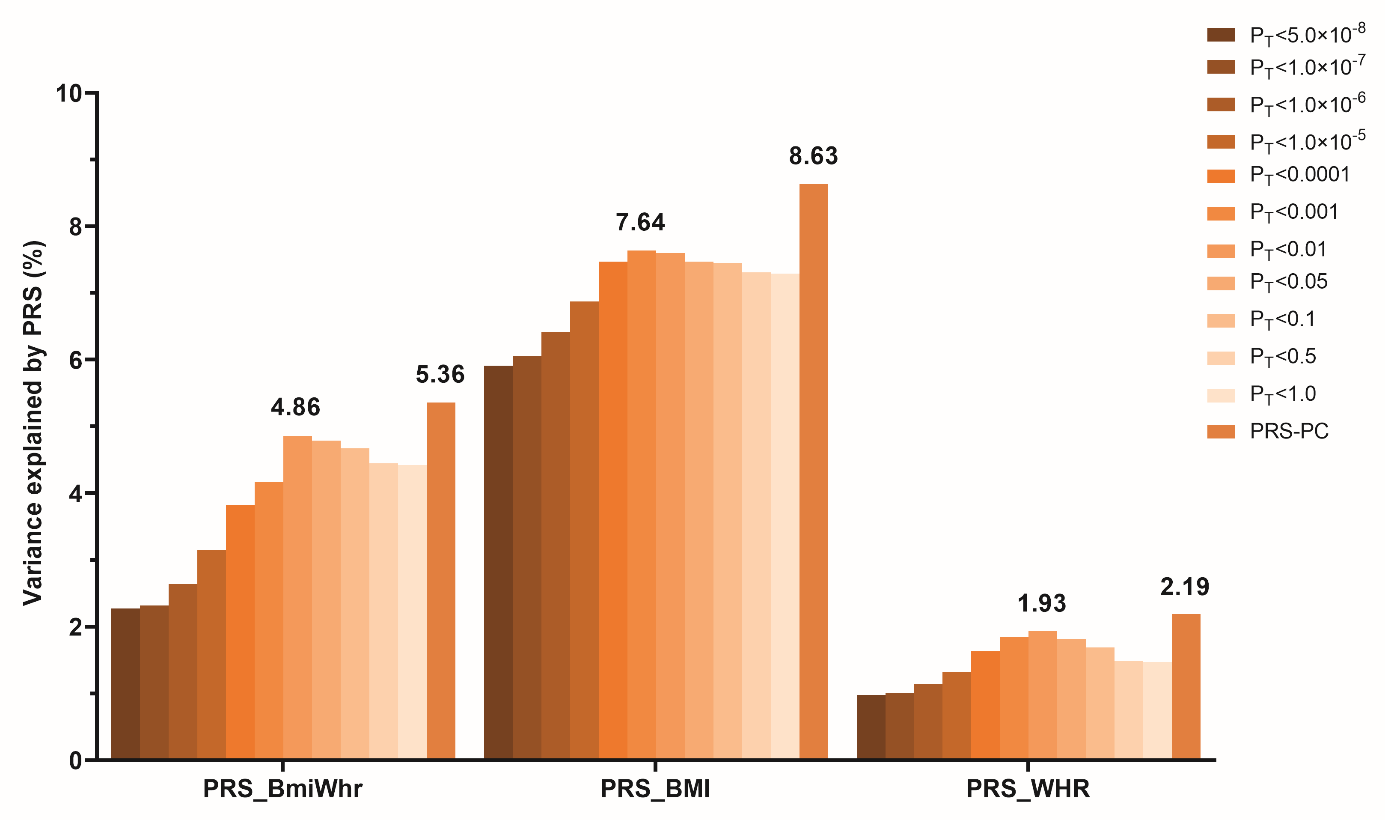

**Figure S5. Variance explained by PRSs for BmiWhr, BMI and WHR** Abbreviations: BmiWhr, the co-occurrence of BMI and WHR; PRS_BmiWhr, the shared polygenic risk score of BMI and WHR; PRS_BMI, polygenic risk score for BMI; PRS_WHR, polygenic risk score for WHR; PRS-PC, the first principal component of PRSs.

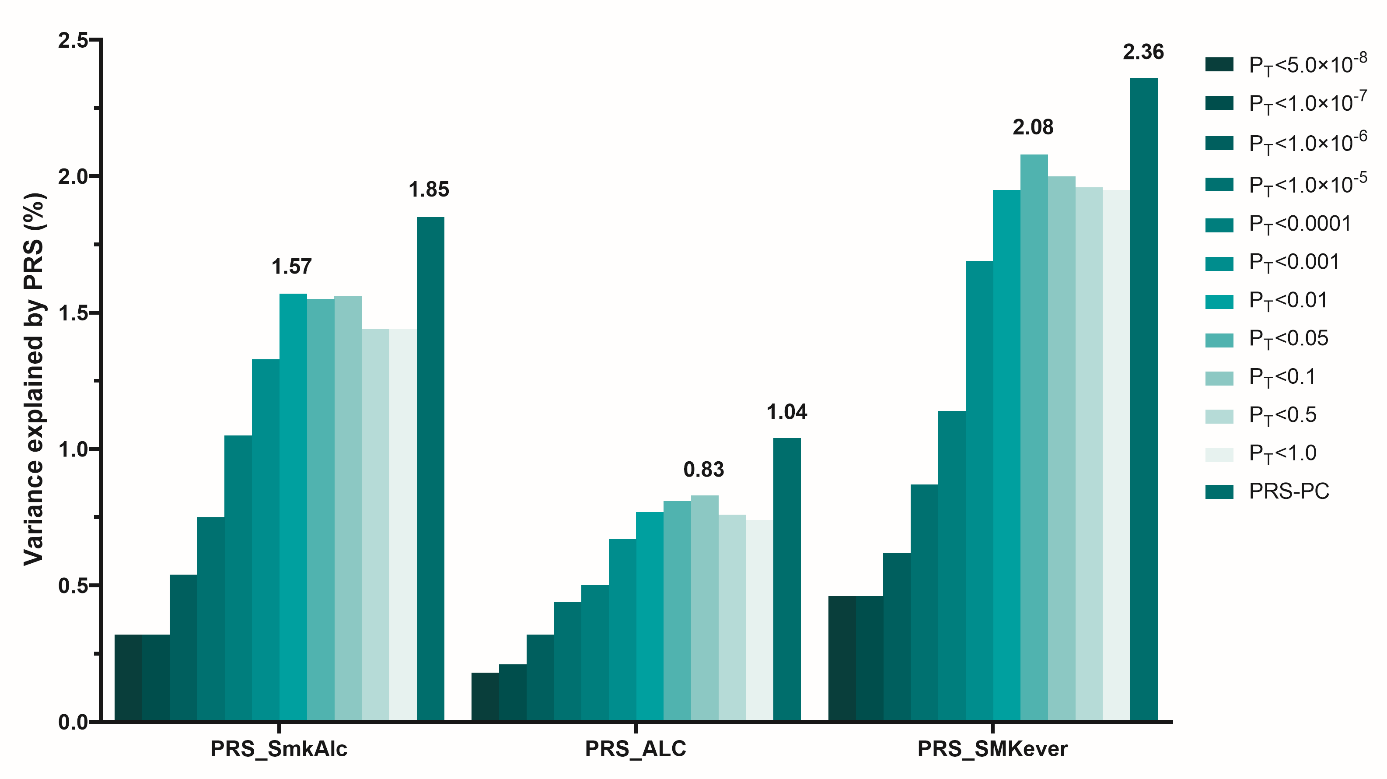

**Figure S6. Variance explained by PRSs for SmkAlc, alcohol use and smoking** Abbreviations: SmkAlc, the co-occurrence of number of cigarettes per day and daily alcohol intake; PRS_SmkAlc, the shared polygenic risk score of smoking and alcohol use; PRS_SMKever, polygenic risk score for smoking; PRS_ALC, polygenic risk score for alcohol use; PRS-PC, the first principal component of PRSs.

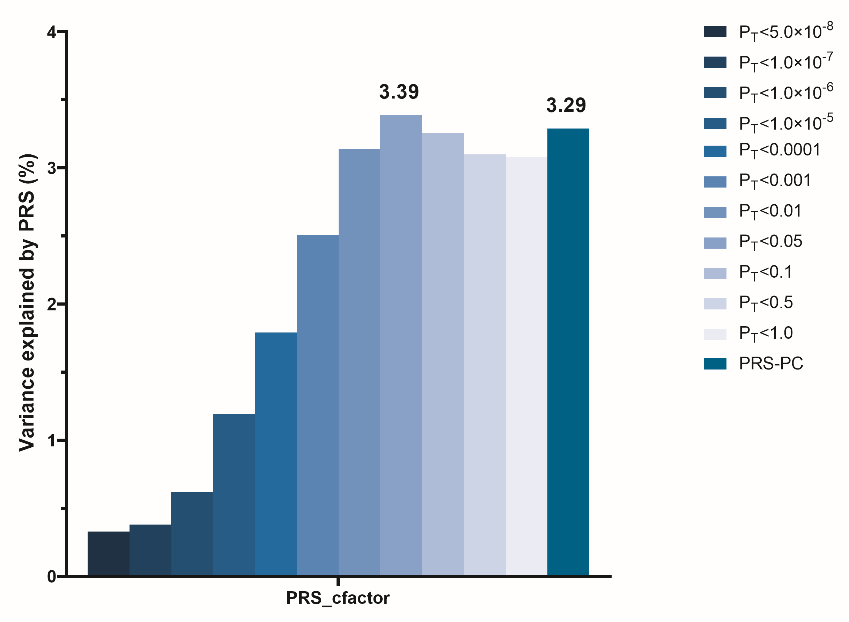

**Figure S7. Variance explained by shared PRSs between three domains for the common factor** Abbreviations: PRS-PC, the first principal component of PRSs. Common factor, the co-occurrence of DepAnx, SmkAlc, and BmiWhr.

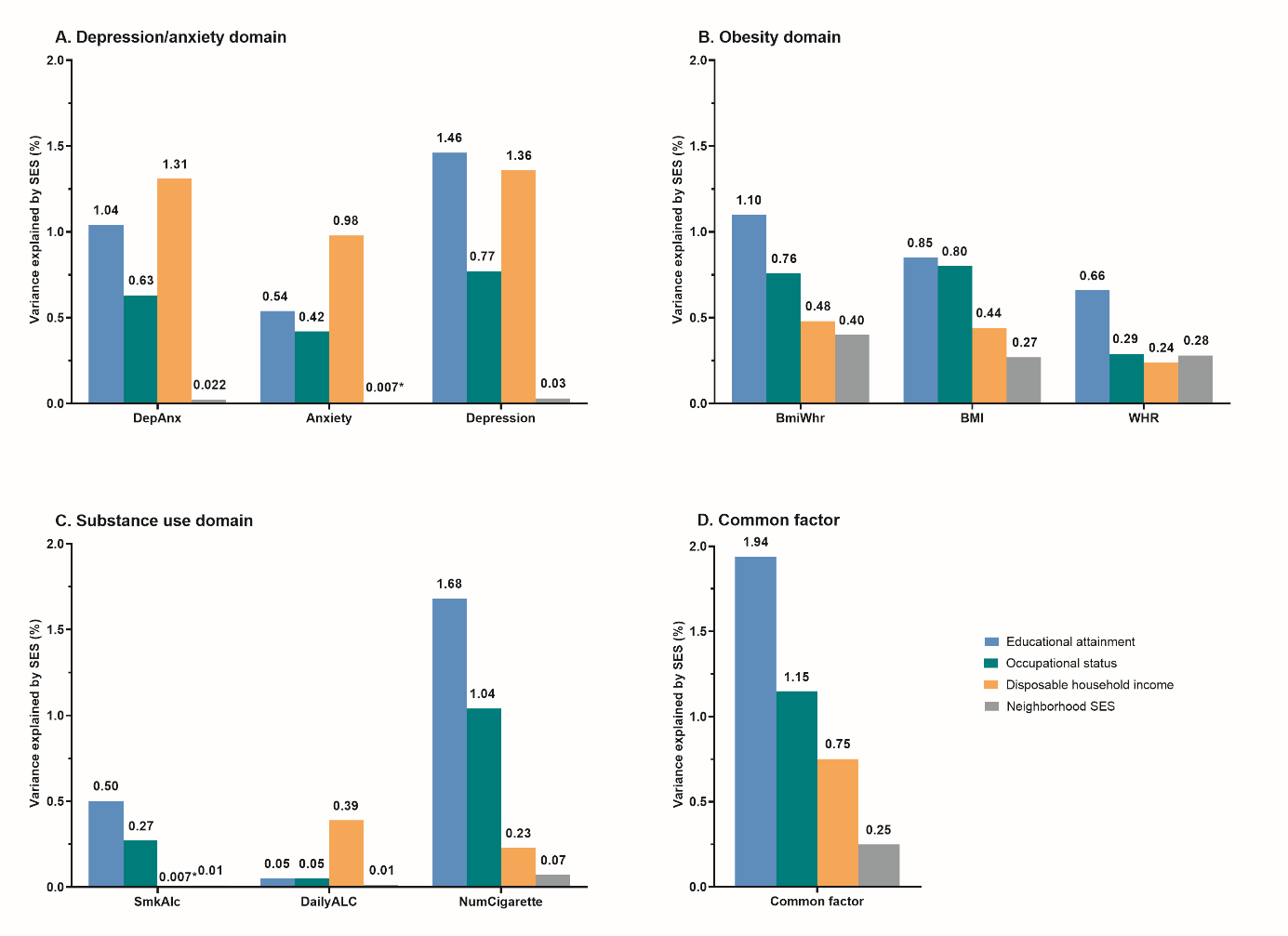

**Figure S8. Variance explained by SES for phenotypes** Abbreviations: BMI, body mass index; WHR, waist-hip-ratio; DailyALC, daily alcohol intake; NumCigarette, number of cigarettes per day; DepAnx, the co-occurrence of depression and anxiety; SmkAlc, the co-occurrence of number of cigarettes per day and daily alcohol intake; BmiWhr, the co-occurrence of BMI and WHR; Common factor, the co-occurrence of DepAnx, SmkAlc, and BmiWhr. * represent p-value non-significant (*p*>0.05).

Socio-economic status moderated the genetic effects on individual outcomes (Figure S9-11). For SES indices at the individual level, lower educational attainment and lower occupational status amplified PRSs effects on depression, anxiety, BMI and smoking (*ps*<0.05), while no interactions between these two SES indices and PRSs were found for WHR and alcohol use. For the household level, lower disposable household income amplified the genetic effects on depression, anxiety and BMI (*ps*<0.05). Inversely, higher disposable household income enlarged the genetic effects on alcohol use (p=0.04). No interactions were found between disposable household income and PRSs for WHR (p=0.95) and smoking (p=0.13). For the neighborhood level, lower NSES amplified the genetic effects on depression and BMI (*ps*<0.05), while no interactions were detected between NSES and PRSs for anxiety, WHR, smoking and alcohol use.

For the depression/anxiety domain, lower educational attainment, lower disposable household income, and lower NSES amplified the shared genetic susceptibility between depression and anxiety on DepAnx (*ps*<0.05) (Figure S9). The interaction between NSES and the shared PRS for DepAnx (p=0.003) was more significant than the interactions for depression (p=0.048) or anxiety (p=0.32) separately. However, although significant interactions between occupational status and PRSs were identified for depression and anxiety separately, no interaction was found between occupational status and the shared PRS for DepAnx (p=0.11). For the obesity domain, lower educational attainment, lower occupational status, and lower disposable household income amplified the shared PRSs effects on BmiWhr (*ps*<0.05), but this was not found for NSES (Figure S10). For the substance use domain, lower educational attainment amplified the shared PRS effect on SmkAlc (p=0.0001), while no interaction effects were detected between the shared PRSs and occupational status, disposable household income and NSES on SmkAlc (Figure S11).

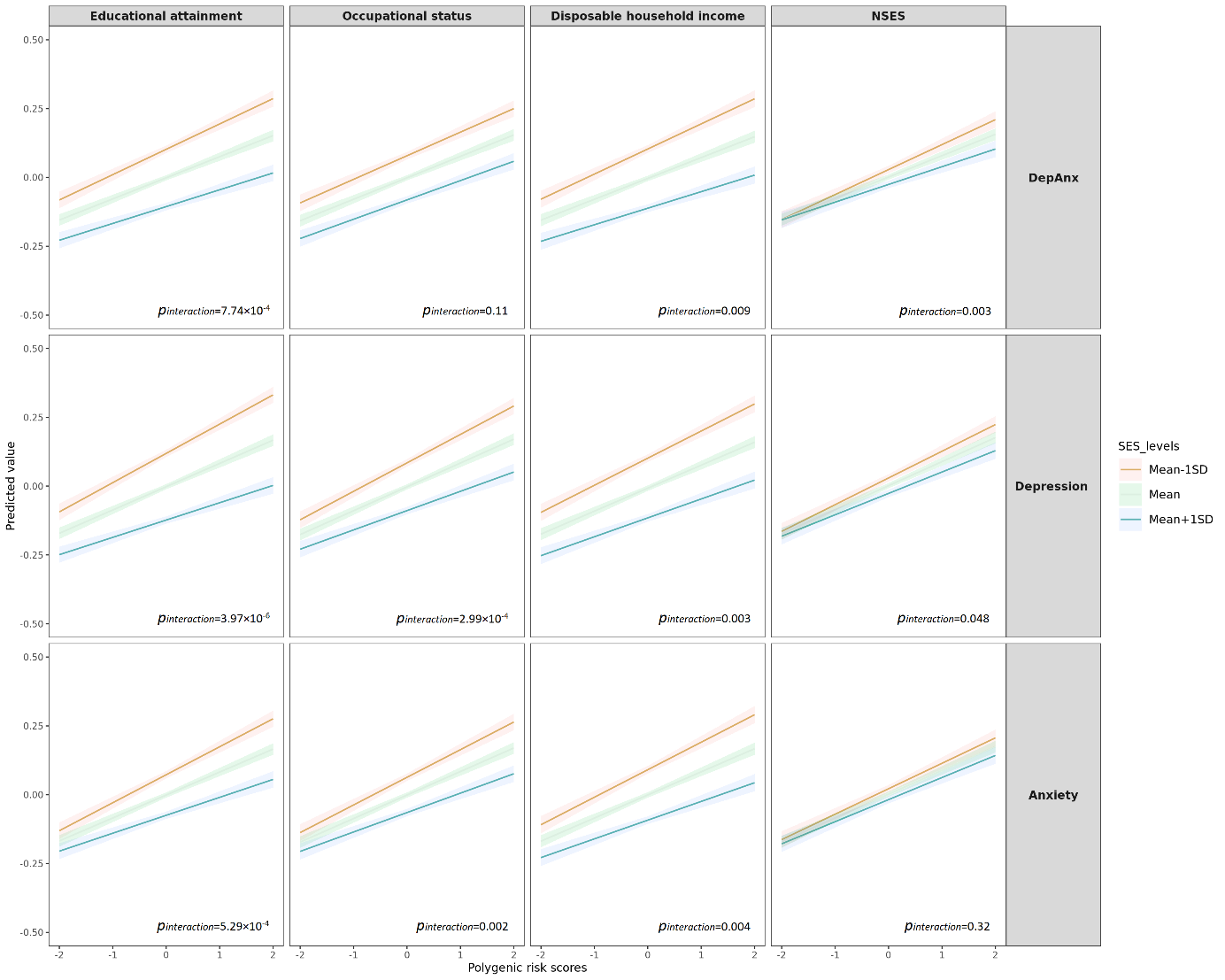

**Figure S9. The interactions between PRSs and SES for depression, anxiety and DepAnx**

Abbreviations: DepAnx, the co-occurrence of depression and anxiety; NSES, neighborhood socio-economic status. Y axis represents the standardized scores of DepAnx, depression and anxiety. Linear mixed regression model adjusted for age, sex, chip (CytoSNP or GSA) and 10 principal components.

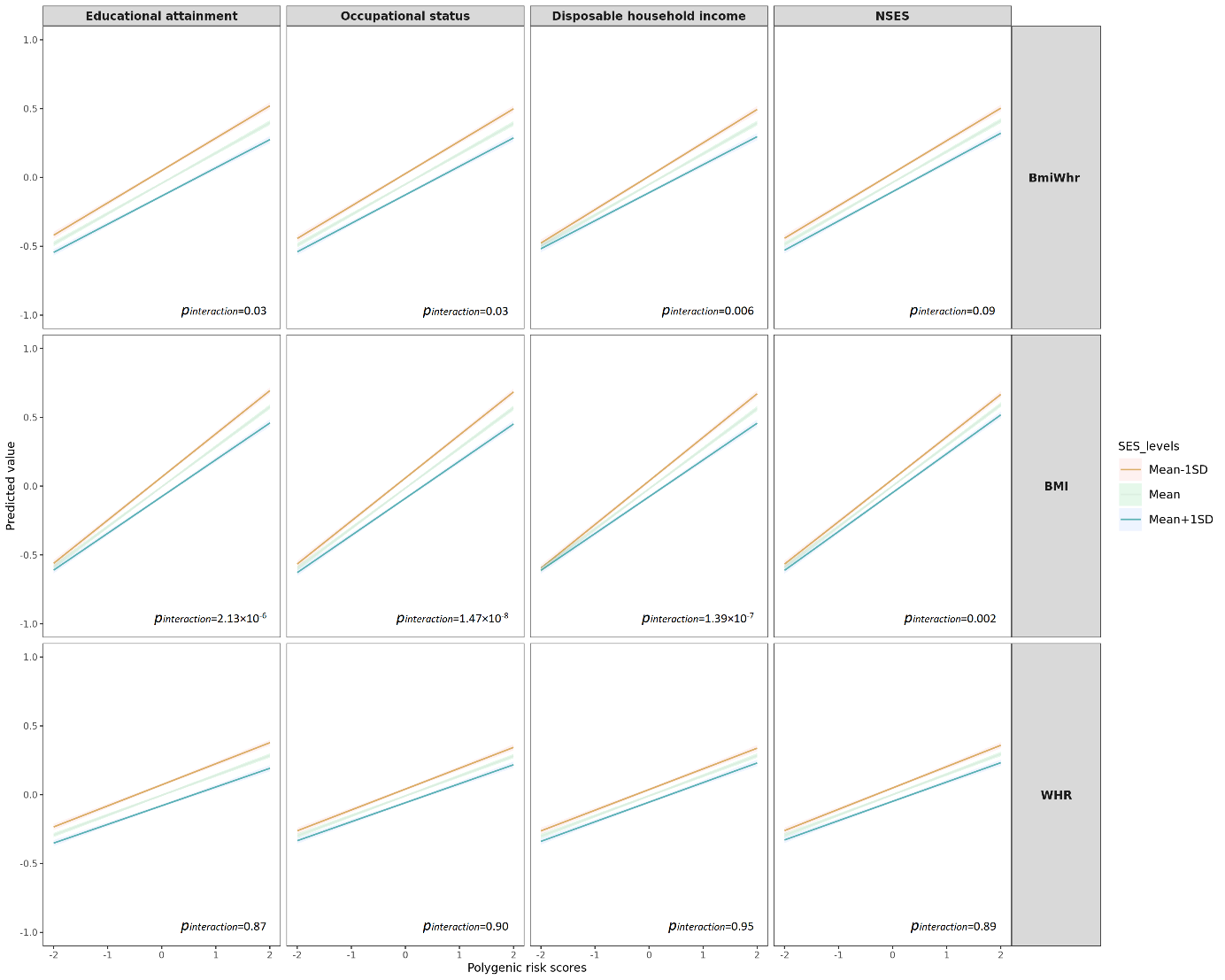

**Figure S10. The interactions between PRSs and SES for BMI, WHR and BmiWhr**

Abbreviations: BmiWhr, the co-occurrence of BMI and WHR; NSES, neighborhood socio-economic status. Y axis represents the standardized scores of BmiWhr, BMI and WHR. Linear mixed regression model adjusted for age, sex, chip (CytoSNP or GSA) and 10 principal components.

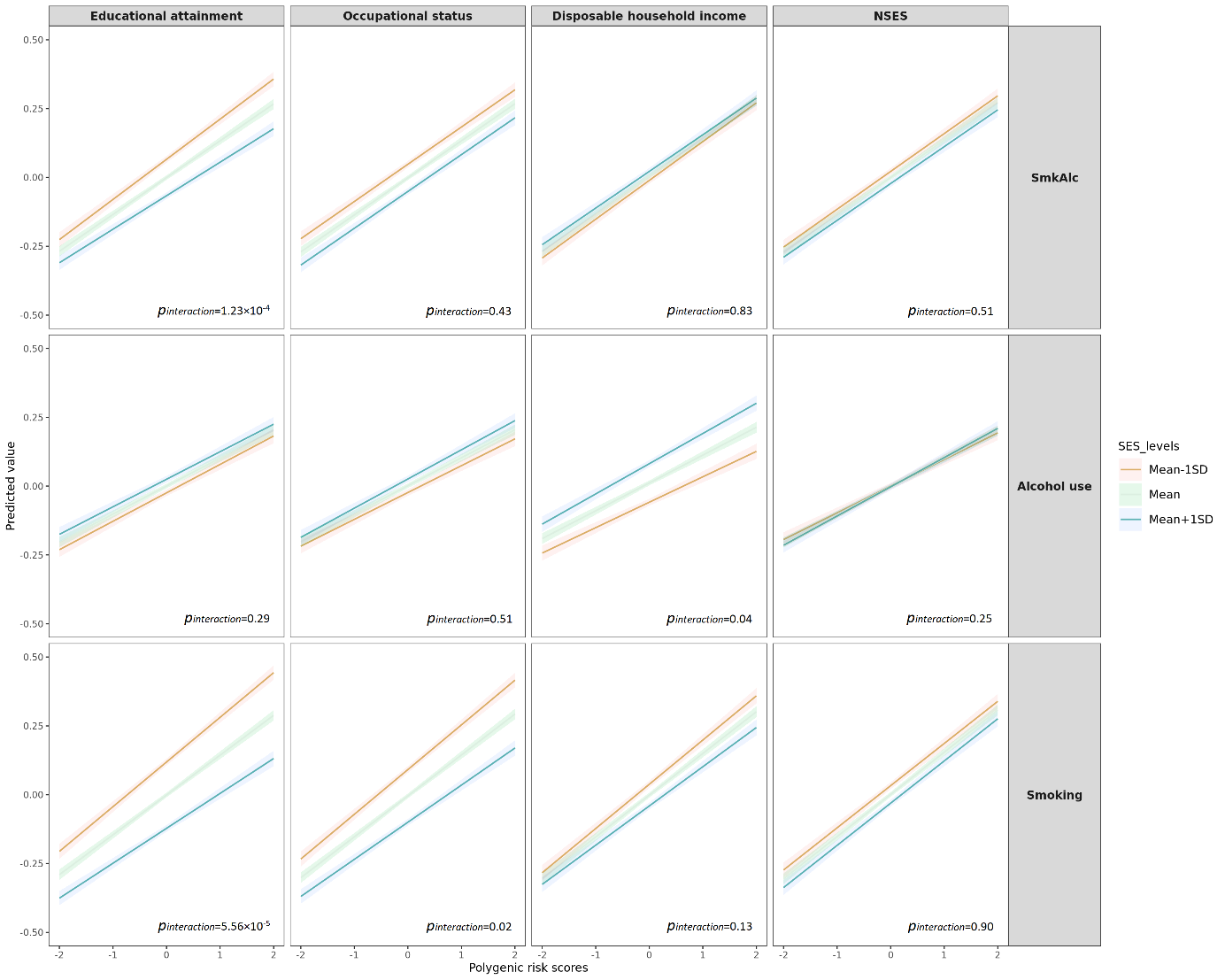

**Figure S11.** **The interactions between PRSs and SES for alcohol use, smoking and SmkAlc** Abbreviations: SmkAlc, the co-occurrence of smoking and alcohol use; NSES, neighborhood socio-economic status. Y axis represents the standardized scores of SmkAlc, alcohol use and smoking. Linear mixed regression model adjusted for age, sex, chip (CytoSNP or GSA) and 10 principal components.

For the shared variance across three disease domains, lower educational attainment (p=0.006) and lower disposable household income (p=0.01) amplified the shared genetic susceptibility across these three domains (Figure S12). No interactions were found between the shared PRS across the three domains with occupational status and NSES.

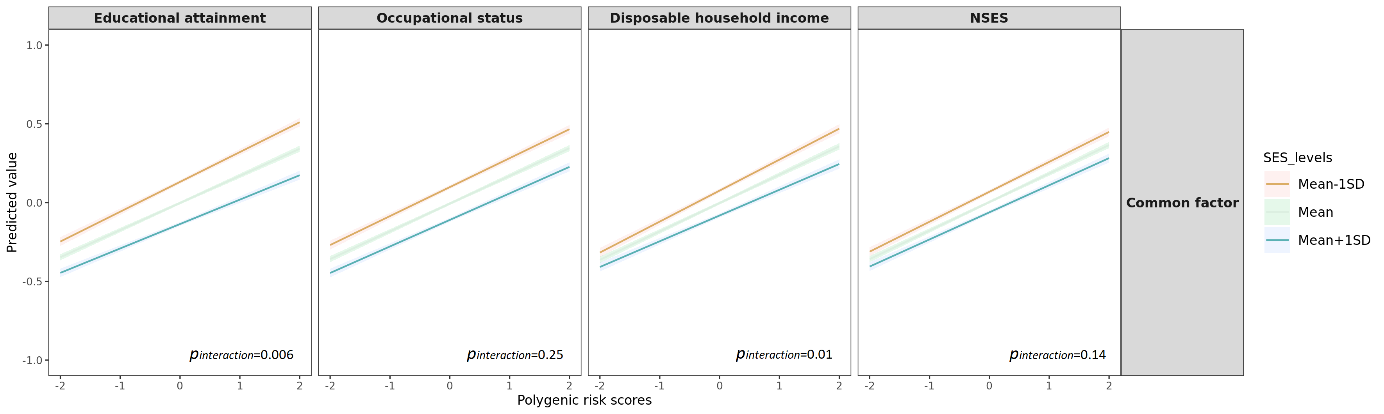

**Figure S12. The interactions between PRSs and SES for the common factor**

Abbreviations: NSES, neighborhood socio-economic status. Common factor, the co-occurrence of depression, anxiety, obesity and substance use. Y axis represents the standardized scores of the common factor. Linear mixed regression model adjusted for age, sex, chip (CytoSNP or GSA) and 10 principal components.

**Table S5. Demographic characteristic of participants with and without missing data at baseline**

| **Variables** | **Analyzed data** | | | **Missing data** | | |
| --- | --- | --- | --- | --- | --- | --- |
|  | **n (%)** | **Age**  **(Mean±SD)** | **Sex (female), n(%)** | **n (%)** | **Age (Mean±SD)** | **Sex (female), n(%)** |
| Outcomes |  |  |  |  |  |  |
| Sum score of depression | 42,669 (84.06) | 41.93±15.45 | 25,103  (58.83) | 8,092 (15.94) | 46.33±15.26 | 4,570  (56.48) |
| Sum score of anxiety | 42,588 (83.90) | 41.88±15.43 | 25,048  (58.81) | 8,173 (16.10) | 46.53±15.32 | 4,625  (56.59) |
| Number of cigarettes per day | 48,603 (95.75) | 42.43±15.24 | 28,818  (59.29) | 2,158 (4.25) | 47.12±19.95 | 855  (39.62) |
| Daily alcohol intake (grams) | 49,930 (98.36) | 42.64±15.23 | 29,221  (58.52) | 831 (1.64) | 41.86±27.25 | 452  (54.39) |
| Body mass index (kg/m2) | 50,628 (99.74) | 42.72±15.42 | 29,594  (58.45) | 133 (0.26) | 9.69±11.91 | 79  (59.40) |
| Waist-hip-ratio | 50,625 (99.73) | 42.72±15.42 | 29,591  (58.45) | 136 (0.27) | 10.31±12.67 | 82  (60.29) |
| Comorbidity |  |  |  |  |  |  |
| DepAnx | 42,318 (83.37) | 41.88±15.44 | 24,877  (58.79) | 8,443 (16.63) | 46.41±15.24 | 4,796  (56.80) |
| SmkAlc | 48,448 (95.44) | 42.36±15.18 | 28,715  (59.27) | 2,313 (4.56) | 48.42±20.26 | 958  (41.42) |
| BmiWhr | 50,625 (99.73) | 42.72±15.42 | 29,591  (58.45) | 136 (0.27) | 10.31±12.67 | 82  (60.29) |
| Common factor | 40,667 (80.11) | 41.74±15.23 | 24,232  (59.59) | 10,094 (19.89) | 46.23±16.06 | 5,441  (53.90) |
| Socio-economic status |  |  |  |  |  |  |
| Educational attainment (years) | 50,204 (98.90) | 42.49±15.45 | 29,349  (58.46) | 557 (1.10) | 55.77±14.35 | 324  (58.17) |
| Occupational status | 48,914 (96.36) | 42.17±15.35 | 28,539  (58.35) | 1,847 (3.64) | 54.73±14.61 | 1,134  (61.40) |
| Disposable household income (euros/month) | 45,214 (89.07) | 42.05±15.46 | 26,095  (57.71) | 5,547 (10.93) | 47.36±15.01 | 3,578  (64.50) |
| Neighborhood socio-economic status | 49,381 (97.28) | 42.62±15.50 | 28,882  (58.49) | 1,380 (2.72) | 43.19±15.72 | 791  (57.32) |
| Latent SES factor | 43,463 (85.62) | 41.56±15.30 | 25,089  (57.72) | 7,298 (14.38) | 49.00±15.14 | 4,584  (62.81) |

Abbreviations: DepAnx, the co-occurrence of depression and anxiety; SmkAlc, the co-occurrence of number of cigarettes per day and daily alcohol intake; BmiWhr, the co-occurrence of BMI and WHR; Common factor, the co-occurrence of DepAnx, SmkAlc, and BmiWhr. Latent SES factor: a latent SES factor combining educational attainment, occupational status, and disposable household income in confirmatory factor analysis.

**Table S6. Attrition analyses between participants with and without missing data**

| **Phenotype** | **Models** | **Total sample** | | | **Sample without missing** | | |
| --- | --- | --- | --- | --- | --- | --- | --- |
|  |  | **n** | ***Beta*** | ***p*** | ***n*** | ***Beta*** | ***p*** |
| Depression | PRS_dep*educational attainment | 42,267 | -0.0315 | 3.97×10^-06^ | 40,306 | -0.0305 | 8.46×10^-06^ |
|  | PRS_dep*occupational status | 41,364 | -0.0249 | 2.99×10^-04^ | 39,474 | -0.0235 | 6.77×10^-04^ |
|  | PRS_dep*disposable household income | 38,357 | -0.0205 | 3.32×10^-03^ | 36,654 | -0.0219 | 1.85×10^-03^ |
|  | PRS_dep*neighborhood SES | 41,551 | -0.0134 | 0.048 | 39,597 | -0.0136 | 0.045 |
| Anxiety | PRS_anx*educational attainment | 42,191 | -0.0374 | 5.29×10^-04^ | 40,306 | -0.0337 | 2.07×10^-03^ |
|  | PRS_anx*occupational status | 41,294 | -0.0332 | 2.23×10^-03^ | 39,474 | -0.0276 | 0.0123 |
|  | PRS_anx*disposable household income | 38,308 | -0.0328 | 3.60×10^-03^ | 36,654 | -0.0319 | 5.23×10^-03^ |
|  | PRS_anx*neighborhood SES | 41,466 | -0.0105 | 0.32 | 39,597 | -0.0086 | 0.43 |
| DepAnx | PRS_depanx*educational attainment | 41,924 | -0.0426 | 7.74×10^-04^ | 40,306 | -0.0456 | 4.22×10^-04^ |
|  | PRS_depanx*occupation status | 41,044 | -0.0206 | 0.11 | 39,474 | -0.0258 | 0.046 |
|  | PRS_depanx*disposable household income | 38,079 | -0.0339 | 9.01×10^-03^ | 36,654 | -0.0347 | 8.76×10^-03^ |
|  | PRS_depanx*neighborhood SES | 41,206 | -0.0372 | 2.97×10^-03^ | 39,597 | -0.0382 | 2.69×10^-03^ |
| BMI | PRS_bmi*educational attainment | 50,072 | -0.0825 | 2.13×10^-06^ | 40,306 | -0.0954 | 9.34×10^-07^ |
|  | PRS_bmi*occupation status | 48,785 | -0.0992 | 1.47×10^-08^ | 39,474 | -0.1283 | 4.88×10^-11^ |
|  | PRS_bmi*disposable household income | 45,092 | -0.0945 | 1.39×10^-07^ | 36,654 | -0.1048 | 1.45×10^-07^ |
|  | PRS_bmi*neighborhood SES | 49,250 | -0.0535 | 2.30×10^-03^ | 39,597 | -0.0528 | 5.97×10^-03^ |
| WHR | PRS_whr*educational attainment | 50,069 | -0.0007 | 0.87 | 40,306 | -0.001 | 0.84 |
|  | PRS_whr*occupational status | 48,782 | -0.0006 | 0.90 | 39,474 | -0.0006 | 0.91 |
|  | PRS_whr*disposable household income | 45,089 | -0.0003 | 0.95 | 36,654 | -0.0004 | 0.95 |
|  | PRS_whr*neighborhood SES | 49,247 | -0.0006 | 0.89 | 39,597 | -0.0005 | 0.91 |
| BmiWhr | PRS_bmiwhr*educational attainment | 50,069 | -0.0099 | 0.03 | 40,306 | -0.0102 | 0.043 |
|  | PRS_bmiwhr*occupational status | 48,782 | -0.0097 | 0.03 | 39,474 | -0.0095 | 0.062 |
|  | PRS_bmiwhr*disposable household income | 45,089 | -0.0130 | 5.70×10^-03^ | 36,654 | -0.0128 | 0.0152 |
|  | PRS_bmiwhr*neighborhood SES | 49,247 | -0.0077 | 0.09 | 39,597 | -0.0081 | 0.108 |
| Smoking | PRS_smk*educational attainment | 48,109 | -0.1255 | 5.56×10^-05^ | 40,306 | -0.1021 | 2.40×10^-03^ |
|  | PRS_smk*occupational status | 46,888 | -0.0720 | 0.02 | 39,474 | -0.0395 | 0.36 |
|  | PRS_smk*disposable household income | 43,422 | -0.0498 | 0.13 | 36,654 | -0.0239 | 0.49 |
|  | PRS_smk*neighborhood SES | 47,285 | 0.0039 | 0.90 | 39,597 | -0.0091 | 0.78 |
| Alcohol use | PRS_alc*educational attainment | 49,404 | -0.0431 | 0.29 | 40,306 | -0.017 | 0.69 |
|  | PRS_alc*occupational status | 48,146 | 0.0268 | 0.51 | 39,474 | 0.0519 | 0.23 |
|  | PRS_alc*disposable household income | 44,685 | 0.0907 | 0.04 | 36,654 | 0.0631 | 0.17 |
|  | PRS_alc*neighborhood SES | 48,574 | 0.0475 | 0.25 | 39,597 | 0.0502 | 0.24 |
| SmkAlc | PRS_smkalc*educational attainment | 47,955 | -0.1088 | 1.23×10^-04^ | 40,306 | -0.0749 | 0.0126 |
|  | PRS_smkalc*occupational status | 46,749 | -0.0226 | 0.43 | 39,474 | 0.007 | 0.81 |
|  | PRS_smkalc*disposable household income | 43,375 | -0.0063 | 0.83 | 36,654 | 0.0201 | 0.52 |
|  | PRS_smkalc*neighborhood SES | 47,137 | -0.0188 | 0.51 | 39,597 | -0.0289 | 0.33 |

Abbreviations: DepAnx, the co-occurrence of depression and anxiety; SmkAlc, the co-occurrence of number of cigarettes per day and daily alcohol intake; BmiWhr, the co-occurrence of BMI and WHR.

**Table S7. Interaction between PRS and latent SES factor for depression, anxiety, BMI, WHR, smoking, alcohol use at individual and aggregated levels**

| **Phenotype** | **Model** | **Beta** | **P** | **R^2^ (%)** | **Model R^2^ (%)** |
| --- | --- | --- | --- | --- | --- |
| Depression | PRS_dep | 0.1151 | < 2.20×10^-16^ | 0.76 | 2.44 |
|  | SES | -0.1242 | < 2.20×10^-16^ | 1.75 |  |
|  | SES*PRS_dep | -0.0187 | 1.19×10^-4^ | 0.038 |  |
| Anxiety | PRS_anx | 0.1882 | < 2.20×10^-16^ | 0.73 | 1.62 |
|  | SES | -0.1433 | < 2.20×10^-16^ | 0.91 |  |
|  | SES*PRS_anx | -0.0309 | 8.04×10^-5^ | 0.040 |  |
| DepAnx | PRS_depanx | 0.1968 | < 2.20×10^-16^ | 0.61 | 2.04 |
|  | SES | -0.2134 | < 2.20×10^-16^ | 1.46 |  |
|  | SES*PRS_depanx | -0.0248 | 6.27×10^-3^ | 0.019 |  |
| BMI | PRS_bmi | 1.298 | < 2.20×10^-16^ | 8.67 | 9.47 |
|  | SES | -0.2622 | < 2.20×10^-16^ | 1.16 |  |
|  | SES*PRS_bmi | -0.0850 | 1.74×10^-11^ | 0.078 |  |
| WHR | PRS_whr | 0.0119 | < 2.20×10^-16^ | 2.14 | 2.63 |
|  | SES | -0.0040 | < 2.20×10^-16^ | 0.61 |  |
|  | SES*PRS_whr | -0.0004 | 0.0664 | 0.005 |  |
| BmiWhr | PRS_ bmiwhr | 0.1392 | < 2.20×10^-16^ | 5.39 | 6.35 |
|  | SES | -0.0406 | < 2.20×10^-16^ | 1.26 |  |
|  | SES*PRS_ bmiwhr | -0.0089 | 4.53×10^-8^ | 0.045 |  |
| Smoking | PRS_ smk | 1.0350 | < 2.20×10^-16^ | 2.36 | 3.84 |
|  | SES | -0.5933 | < 2.20×10^-16^ | 1.74 |  |
|  | SES*PRS_ smk | -0.0970 | 1.33×10^-5^ | 0.040 |  |
| Alcohol use | PRS_alc | 0.9750 | < 2.20×10^-16^ | 1.03 | 1.20 |
|  | SES | 0.2654 | < 2.20×10^-16^ | 0.18 |  |
|  | SES*PRS_alc | 0.0127 | 0.6662 | 3.81×10^-4^ |  |
| SmkAlc | PRS_smkalc | 0.9016 | < 2.20×10^-16^ | 1.86 | 2.18 |
|  | SES | -0.2543 | < 2.20×10^-16^ | 0.34 |  |
|  | SES*PRS_smkalc | -0.0516 | 0.0112 | 0.013 |  |
| Common factor | PRS_cfactor | 0.1393 | < 2.20×10^-16^ | 3.23 | 5.09 |
|  | SES | -0.0760 | < 2.20×10^-16^ | 2.16 |  |
|  | SES*PRS_cfactor | -0.0095 | 2.55×10^-4^ | 0.029 |  |

Abbreviations: DepAnx, the co-occurrence of depression and anxiety; SmkAlc, the co-occurrence of number of cigarettes per day and daily alcohol intake; BmiWhr, the co-occurrence of BMI and WHR; Common factor, the co-occurrence of DepAnx, SmkAlc, and BmiWhr. SES: a latent SES factor combining educational attainment, occupational status, and disposable household income in confirmatory factor analysis.

**Table S8. Interactions between PRSs and socio-economic status for different scales of phenotypes**

| **Phenotype** | **Models** | **n** | **Original scales** | | **Inverse normal transformation of residuals** | |
| --- | --- | --- | --- | --- | --- | --- |
|  |  |  | ***Beta*** | ***p*** | ***Beta*** | ***p*** |
| Depression | PRS_dep*educational attainment | 42,267 | -0.0315 | 3.97×10^-06^ | -0.0112 | 0.02 |
|  | PRS_dep*occupational status | 41,364 | -0.0249 | 2.99×10^-04^ | -0.0095 | 0.06 |
|  | PRS_dep*disposable household income | 38,357 | -0.0205 | 3.32×10^-03^ | -0.0110 | 0.03 |
|  | PRS_dep*neighborhood SES | 41,551 | -0.0134 | 0.048 | -0.0097 | 0.048 |
| Anxiety | PRS_anx*educational attainment | 42,191 | -0.0374 | 5.29×10^-04^ | -0.0166 | 8.28×10^-04^ |
|  | PRS_anx*occupational status | 41,294 | -0.0332 | 2.23×10^-03^ | -0.0131 | 9.22×10^-03^ |
|  | PRS_anx*disposable household income | 38,308 | -0.0328 | 3.60×10^-03^ | -0.0142 | 6.48×10^-03^ |
|  | PRS_anx*neighborhood SES | 41,466 | -0.0105 | 0.32 | -0.0027 | 0.59 |
| DepAnx | PRS_depanx*educational attainment | 41,924 | -0.0426 | 7.74×10^-04^ | -0.0078 | 0.12 |
|  | PRS_depanx*occupation status | 41,044 | -0.0206 | 0.11 | -0.0030 | 0.55 |
|  | PRS_depanx*disposable household income | 38,079 | -0.0339 | 9.01×10^-03^ | -0.0099 | 0.06 |
|  | PRS_depanx*neighborhood SES | 41,206 | -0.0372 | 2.97×10^-03^ | -0.0080 | 0.11 |
| BMI | PRS_bmi*educational attainment | 50,072 | -0.0825 | 2.13×10^-06^ | -0.0156 | 5.15×10^-04^ |
|  | PRS_bmi*occupation status | 48,785 | -0.0992 | 1.47×10^-08^ | -0.0128 | 5.00×10^-03^ |
|  | PRS_bmi*disposable household income | 45,092 | -0.0945 | 1.39×10^-07^ | -0.0155 | 9.20×10^-04^ |
|  | PRS_bmi*neighborhood SES | 49,250 | -0.0535 | 2.30×10^-03^ | -0.0088 | 0.054 |
| WHR | PRS_whr*educational attainment | 50,069 | -0.0007 | 0.87 | -0.0105 | 0.02 |
|  | PRS_whr*occupational status | 48,782 | -0.0006 | 0.90 | -0.0084 | 0.07 |
|  | PRS_whr*disposable household income | 45,089 | -0.0003 | 0.95 | -0.0046 | 0.33 |
|  | PRS_whr*neighborhood SES | 49,247 | -0.0006 | 0.89 | -0.0091 | 0.048 |
| BmiWhr | PRS_bmiwhr*educational attainment | 50,069 | -0.0099 | 0.03 | -0.0132 | 3.10×10^-03^ |
|  | PRS_bmiwhr*occupational status | 48,782 | -0.0097 | 0.03 | -0.0138 | 2.60×10^-03^ |
|  | PRS_bmiwhr*disposable household income | 45,089 | -0.0130 | 5.70×10^-03^ | -0.0198 | 2.71×10^-05^ |
|  | PRS_bmiwhr*neighborhood SES | 49,247 | -0.0077 | 0.09 | -0.0113 | 0.01 |
| Smoking | PRS_smk*educational attainment | 48,109 | -0.1255 | 5.56×10^-05^ | -0.0171 | 2.07×10^-04^ |
|  | PRS_smk*occupational status | 46,888 | -0.0720 | 0.02 | -0.0104 | 0.02 |
|  | PRS_smk*disposable household income | 43,422 | -0.0498 | 0.13 | -0.0037 | 0.44 |
|  | PRS_smk*neighborhood SES | 47,285 | 0.0039 | 0.90 | 0.0053 | 0.25 |
| Alcohol use | PRS_alc*educational attainment | 49,404 | -0.0431 | 0.29 | -0.0008 | 0.86 |
|  | PRS_alc*occupational status | 48,146 | 0.0268 | 0.51 | 0.0068 | 0.14 |
|  | PRS_alc*disposable household income | 44,685 | 0.0907 | 0.04 | 0.0063 | 0.20 |
|  | PRS_alc*neighborhood SES | 48,574 | 0.0475 | 0.25 | 0.0104 | 0.03 |
| SmkAlc | PRS_smkalc*educational attainment | 47,955 | -0.1088 | 1.23×10^-04^ | -0.0088 | 0.06 |
|  | PRS_smkalc*occupational status | 46,749 | -0.0226 | 0.43 | 0.0036 | 0.44 |
|  | PRS_smkalc*disposable household income | 43,375 | -0.0063 | 0.83 | -0.0016 | 0.74 |
|  | PRS_smkalc*neighborhood SES | 47,137 | -0.0188 | 0.51 | 0.0018 | 0.70 |
| Common factor | PRS_cfactor*educational attainment | 40,306 | -0.0139 | 5.90×10^-03^ | -0.0008 | 0.87 |
|  | PRS_cfactor*occupational status | 39,474 | -0.0059 | 0.25 | 0.0046 | 0.37 |
|  | PRS_cfactor*disposable household income | 36,654 | -0.0134 | 0.01 | 0.0022 | 0.68 |
|  | PRS_cfactor*neighborhood SES | 39,597 | -0.0073 | 0.14 | 0.0045 | 0.37 |

Abbreviations: DepAnx, the co-occurrence of depression and anxiety; SmkAlc, the co-occurrence of number of cigarettes per day and daily alcohol intake; BmiWhr, the co-occurrence of BMI and WHR; Common factor, the co-occurrence of DepAnx, SmkAlc, and BmiWhr. Residuals were generated from the regression model adjusting for age, sex, chip (CytoSNP or GSA) and 10 principal components. After changing the outcome scales into inverse normal transformation of residuals, 9 of 40 interactions became non-siginificant, 4 of 40 interactions became significant.

**Table S9. Pearson correlations between PRSs and socio-economic status**

| **Phenotype** | **PRS** | **SES** | **Correlation** | ***p*** |
| --- | --- | --- | --- | --- |
| Depression | PRS_dep | Educational attainment | -0.040 | < 2.20×10^-16^ |
|  | PRS_dep | Occupational status | -0.027 | 2.61×10^-09^ |
|  | PRS_dep | Disposable household income | -0.025 | 8.71×10^-08^ |
|  | PRS_dep | Neighborhood SES | -0.015 | 6.57×10^-04^ |
| Anxiety | PRS_anx | Educational attainment | -0.032 | 3.33×10^-13^ |
|  | PRS_anx | Occupational status | -0.020 | 1.41×10^-05^ |
|  | PRS_anx | Disposable household income | -0.018 | 1.51×10^-04^ |
|  | PRS_anx | Neighborhood SES | -0.023 | 1.89×10^-07^ |
| DepAnx | PRS_depanx | Educational attainment | -0.023 | 1.53×10^-07^ |
|  | PRS_depanx | Occupational status | -0.013 | 0.0041 |
|  | PRS_depanx | Disposable household income | -0.018 | 1.73×10^-04^ |
|  | PRS_depanx | Neighborhood SES | -0.012 | 0.0099 |
| BMI | PRS_bmi | Educational attainment | -0.075 | < 2.20×10^-16^ |
|  | PRS_bmi | Occupational status | -0.055 | < 2.20×10^-16^ |
|  | PRS_bmi | Disposable household income | -0.029 | 4.63×10^-10^ |
|  | PRS_bmi | Neighborhood SES | -0.026 | 1.38×10^-08^ |
| WHR | PRS_whr | Educational attainment | -0.053 | < 2.20×10^-16^ |
|  | PRS_whr | Occupational status | -0.041 | < 2.20×10^-16^ |
|  | PRS_whr | Disposable household income | -0.036 | 3.10×10^-14^ |
|  | PRS_whr | Neighborhood SES | -0.017 | 1.83×10^-04^ |
| BmiWhr | PRS_bmiwhr | Educational attainment | -0.068 | < 2.20×10^-16^ |
|  | PRS_bmiwhr | Occupational status | -0.052 | < 2.20×10^-16^ |
|  | PRS_bmiwhr | Disposable household income | -0.033 | 1.11×10^-12^ |
|  | PRS_bmiwhr | Neighborhood SES | -0.025 | 2.07×10^-08^ |
| Smoking | PRS_smk | Educational attainment | -0.088 | < 2.20×10^-16^ |
|  | PRS_smk | Occupational status | -0.056 | < 2.20×10^-16^ |
|  | PRS_smk | Disposable household income | -0.037 | 4.14×10^-15^ |
|  | PRS_smk | Neighborhood SES | -0.022 | 9.71×10^-07^ |
| Alcohol use | PRS_alc | Educational attainment | 0.010 | 0.0303 |
|  | PRS_alc | Occupational status | 0.010 | 0.0252 |
|  | PRS_alc | Disposable household income | 0.015 | 0.0010 |
|  | PRS_alc | Neighborhood SES | 0.002 | 0.6606 |
| SmkAlc | PRS_smkalc | Educational attainment | -0.030 | 2.42×10^-11^ |
|  | PRS_smkalc | Occupational status | -0.014 | 0.0028 |
|  | PRS_smkalc | Disposable household income | -0.002 | 0.6532 |
|  | PRS_smkalc | Neighborhood SES | -0.008 | 0.0842 |
| Common factor | PRS_cfactor | Educational attainment | -0.071 | < 2.20×10^-16^ |
|  | PRS_cfactor | Occupational status | -0.043 | < 2.20×10^-16^ |
|  | PRS_cfactor | Disposable household income | -0.025 | 1.62×10^-07^ |
|  | PRS_cfactor | Neighborhood SES | -0.022 | 1.39×10^-06^ |

Abbreviations: DepAnx, the co-occurrence of depression and anxiety; SmkAlc, the co-occurrence of number of cigarettes per day and daily alcohol intake; BmiWhr, the co-occurrence of BMI and WHR; Common factor, the co-occurrence of DepAnx, SmkAlc, and BmiWhr.

**Table S10. Genetic and phenotypic correlations between outcomes**

| **Phenotype1** | **Phenotype2** | **Genetic correlation** | | **Phenotypic correlation** | |
| --- | --- | --- | --- | --- | --- |
|  |  | ***rG*** | ***se*** | ***rP*** | ***se*** |
| Depression | Anxiety | 0.90 | 0.05 | 0.67 | 0.003 |
| BMI | WHR | 0.61 | 0.03 | 0.37 | 0.004 |
| NumCigarette | DailyALC | 0.41 | 0.02 | 0.26 | 0.004 |
| DepAnx | BmiWhr | 0.16 | 0.03 | 0.02 | 0.005 |
| DepAnx | SmkAlc | 0.20 | 0.03 | 0.06 | 0.005 |
| BmiWhr | SmkAlc | 0.14 | 0.02 | 0.25 | 0.004 |

Abbreviations: BMI, body mass index; WHR, waist-hip-ratio; DailyALC, daily alcohol intake; NumCigarette, number of cigarettes per day; DepAnx, the co-occurrence of depression and anxiety; SmkAlc, the co-occurrence of number of cigarettes per day and daily alcohol intake; BmiWhr, the co-occurrence of BMI and WHR. Genetic correlation was estimated using the summary statistics from GWAS in LD Score regression. Phenotypic correlation was estimated using Pearson correlation. All *rG* and *rP* are significant (p<0.05).

In Genomic SEM, Q_SNP_ is a ꭓ^2^-distributed test statistics, which test the heterogeneity of effect sizes of each SNPs across each individual phenotype and the common factor [12]. High heterogeneity of SNP effects was defined as Q_SNP_ P-value<5×10^-8^. Low heterogeneity of SNP effects was defined as Q_SNP_ P-value>0.005. A more strict low heterogeneity of SNP effects was defined as Q_SNP_ P-value>0.05. PRSs were calculated for total SNPs, low heterogeneity SNPs, strict low heterogeneity SNPs, and high heterogeneity SNPs. The number of SNPs included in different PRSs were shown in Table S11. The variance explained by these 4 PRSs for common factor was shown in Figure S13. The variance explained by PRS of low heterogeneity SNPs (3.11% and 3.28%) for common factor is similar with that of total PRS (3.29%).

**Table S11. The number of SNPs with different heterogeneity in Q_SNP_**

| **PRS** | **SNPs** | **Independent SNPs** |
| --- | --- | --- |
| PRS_all SNPs | 5,234,508 | 144,876 |
| PRS_Q_SNPs__low_p>0.005 | 5,020,929 | 144,188 |
| PRS_Q_SNPs__low_p>0.05 | 4,509,951 | 140,506 |
| PRS_Q_SNPs__high_p<5×10^-8^ | 3,340 | 144 |

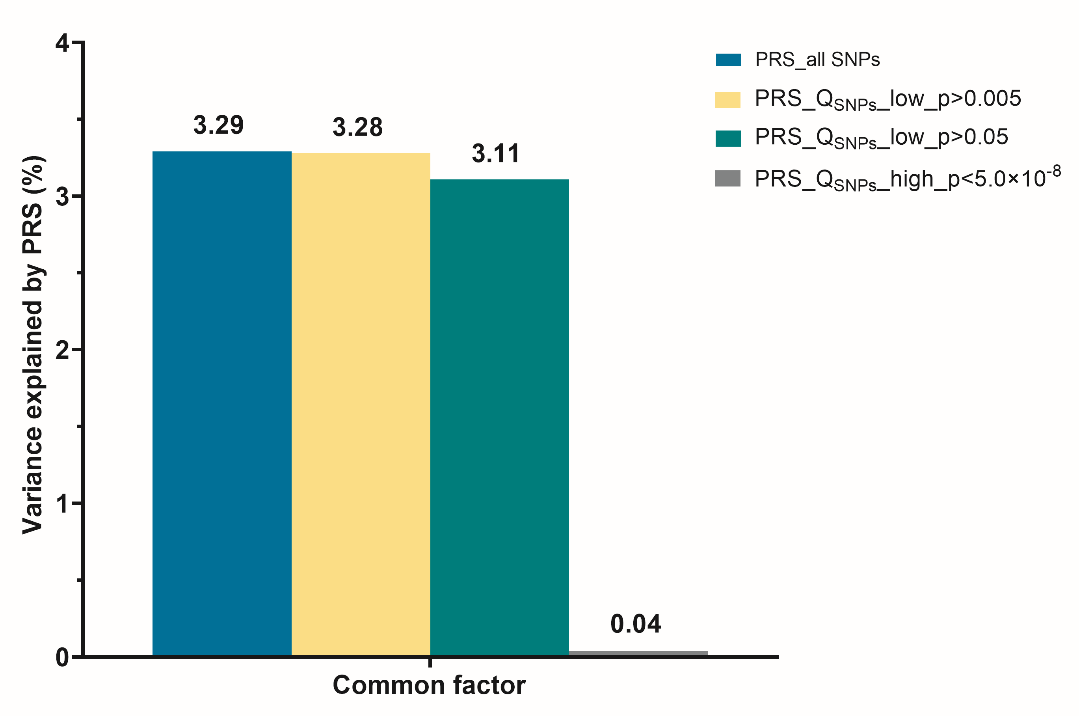

**Figure S13. Variance explained by Q_SNP_ test PRSs for common factor** Abbreviations: PRS, polygenic risk score; PRS_Q_SNPs__low_p>0.005: PRS was calculated based on low heterogeneity SNPs in Q_SNP_ (p>0.005); PRS_Q_SNPs__low_p>0.05: PRS was calculated based on more strict low heterogeneity SNPs in Q_SNP_ (p>0.05); PRS_Q_SNPs__high_p<5×10^-8^: PRS was calculated based on high heterogeneity SNPs in Q_SNP_ (p>5×10^-8^).

**UMCG Genetics Lifelines Initiative (UGLI) group author**

Lifelines Cohort Study

Raul Aguirre-Gamboa (1), Patrick Deelen (1), Lude Franke (1), Jan A Kuivenhoven (2), Esteban A Lopera Maya (1), Ilja M Nolte (3), Serena Sanna (1), Harold Snieder (3), Morris A Swertz (1), Peter M. Visscher (3,4), Judith M Vonk (3), Cisca Wijmenga (1)

(1) Department of Genetics, University of Groningen, University Medical Center Groningen, The Netherlands
(2) Department of Pediatrics, University of Groningen, University Medical Center Groningen, The Netherlands
(3) Department of Epidemiology, University of Groningen, University Medical Center Groningen, The Netherlands
(4) Institute for Molecular Bioscience, The University of Queensland, Brisbane, Queensland, Australia.
